# Quantification of T- and B-cell immune receptor distribution diversity characterizes immune cell infiltration and lymphocyte heterogeneity in clear cell renal cell carcinoma

**DOI:** 10.1101/2021.06.15.21258987

**Authors:** Meghan C. Ferrall-Fairbanks, Nicholas Chakiryan, Boris I. Chobrutskiy, Youngchul Kim, Jamie K. Teer, Anders Berglund, James J. Mulé, Michelle Fournier, Erin M. Siegel, Jasreman Dhillon, Seyed Shayan A. Falasiri, Juan F. Arturo, Esther N. Katende, George Blanck, Brandon J. Manley, Philipp M. Altrock

## Abstract

Immune-modulating systemic therapies are often used to treat advanced cancer such as metastatic clear cell renal cell carcinoma (ccRCC). Used alone, sequence-based biomarkers neither accurately capture patient dynamics nor the tumor immune microenvironment. To better understand the tumor ecology of this immune microenvironment, we quantified tumor infiltration across two distinct ccRCC patient tumor cohorts using complementarity determining region-3 (CDR3) sequence recovery counts in tumor-infiltrating lymphocytes and a generalized diversity index (GDI) for CDR3 sequence distributions. GDI can be understood as a curve over a continuum of diversity scales which allows sensitive characterization of distributions to capture sample richness, evenness, and subsampling uncertainty, along with other important metrics that characterize tumor heterogeneity. For example, richness quantified the total unique sequence count, while evenness quantified similarities across sequence frequencies. Significant differences in receptor sequence diversity across gender and race revealed that patients with larger and more clinically aggressive tumors had increased richness of recovered tumoral CDR3 sequences, specifically in those from T-cell receptor alpha and B-cell immunoglobulin lambda light chain. The GDI inflection point (IP) allowed for a novel and robust measure of distribution evenness. High IP values associated with improved overall survival, suggesting that normal-like sequence distributions lead to better outcomes. These results propose a new quantitative tool that can be used to better characterize patient-specific differences related to immune cell infiltration, and to identify unique characteristics of tumor-infiltrating lymphocyte heterogeneity in ccRCC and other malignancies.

## INTRODUCTION

Renal cell carcinoma ranks seventh and tenth among the most diagnosed cancers among men and women in the US, respectively, accounting for 3.8% of all cancer cases and 2.5% of all cancer deaths (1). The most common type of renal cell carcinoma is clear cell renal cell carcinoma (ccRCC). Historically, metastatic ccRCC has been one of the first malignancies successfully treated with immune-modulating systemic therapy, using interleukin-2 and interferon-α (2). Immune checkpoint inhibitors (ICI) such as nivolumab, ipilimumab, pembrolizumab, and avelumab, have emerged as the first line therapy for metastatic ccRCC, typically administered in combination with each other or with a targeted therapeutic agent (3). The arrival of immune checkpoint inhibitors has precipitated a tremendous research effort aiming to accurately characterize the tumor-immune microenvironment and explore potential biomarkers to predict ICI response, for which robust markers have been largely elusive. Most investigations have focused on tumor-centric variables including somatic mutations and gene expression. Fewer studies have been focused on host factors that contribute to the microenvironment or focused on how differences among these factors may affect clinical or therapeutic outcomes.

Across tumor types, response to ICI has been correlated with higher frequencies of somatic mutations that are believed to give rise to tumor-specific neoantigens, and to stimulate a robust antitumor immune response (4–6). In contrast, analyses of renal cell carcinomas have demonstrated a relatively low frequency of somatic mutations, yet very high levels of immune cell infiltration. These findings suggest that a high mutational burden is not solely responsible for inducing immune infiltration in ccRCC (7–11). Additionally, recent work has demonstrated that CD8+ T cell infiltration alone does not predict response to ICI. Refinements of characterizing immune cell populations are needed to understand the microenvironment and the biology underlying ICI response (12).

To initiate an anti-tumor immune response, tumor-specific neoantigens first require recognition by a T- or B-cell receptor (TCR, BCR) on a tumor infiltrating lymphocyte. The tumor infiltrating lymphocyte, complementarity determining region-3 (CDR3) is a highly variable region in the TCR/BCR that provides a complementary binding surface for antigens and largely determines the antigen specificity of the receptor. Investigations have shown the promise of CDR3-features as prognostic biomarkers for several malignancies (13–16), using sequencing and bioinformatic pipelines to recover single reads that represent the CDR3 amino acid sequence. These reads can be quantified as total recovered reads, potentially as a primary metric of immune infiltration (14–16). However, total count-based measures of CDR3 variability are unlikely to reflect the underlying complex biology of host adaptive immune response with the same accuracy as other measures of receptor diversity (17–19).

We hypothesized that immune cell receptor sequence diversity recapitulates important features of tumor biology such as origin, environment-driven evolution, and progression risk. We leverage properties of a generalized diversity index (GDI) (20–22), a measure applied in ecology and evolution, to quantify CDR3 diversity, and assess whether this diversity is associated with important clinicopathologic outcomes in ccRCC. GDI is evaluated as a continuous function along a range of order of diversity (q) values (20–22). At low values of q (low-q GDI), the index is a measure of distribution richness, i.e., the count of distinct types, sequences or clones, while the value at q=1 is closely related to Shannon’s diversity index (23). At high values of q (high-q GDI), the index approaches a measure of evenness or dominance, i.e., focusing on the dominant clone or sequence and its frequency. Here, we applied these diversity metrics to ccRCC tumor samples, and assessed the properties of GDI and their utility to serve as possible prognostic markers. We assessed tumor infiltrating lymphocyte TCR and BCR CDR3 diversity across the entire range of q, and for isolated values that have direct statistical interpretations. We analyzed two independent cohorts of ccRCC patients with bulk RNA-sequencing samples; the Moffitt Total Cancer Care (TCC) cohort (24), and the Clinical Proteomic Tumor Analysis Consortium 3 (CPTAC-3) cohort (25).

## MATERIALS AND METHODS

### Clinical samples

Following institutional review board approval (H. Lee Moffitt Cancer Center’s Total Cancer Care protocol MCC# 14690; approved by the institutional review board; Advarra IRB Pro00014441), we retrospectively obtained clinicopathological and bulk RNA-sequencing patient data from electronic medical records, where all patients had provided written consent under the institutional Total Cancer Care (TCC) Protocol. RNA was prepared using the Qiagen (Venlo, NL) RNAeasy plus mini kit for RNA (frozen tissue) or the Qiagen All prep FFPE DNA/RNA kit (FFPE tissue). RNAseq sequencing libraries were prepared using the standard Illumina TruSeq RNA Access kit (now called TruSeq RNA Exome), according to manufacturer protocols. RNAseq libraries were sequenced on an Illumina HiSeq 4000 according to manufacturer protocols. RNAseq reads were aligned to the human reference genome (hs37d5) in an intron-aware manner with STAR (26). Table 1 shows a summary of the clinical information obtained from individuals in the Moffitt TCC Cohort. Relevant clinical and pathological outcomes available from the Moffitt TCC Cohort, including ranges of percentage of tumor with EGFR spice variant alpha are recorded in Table 1. Summaries of numbers of reads per samples in each of the cohorts is available in Supplemental Figure S1.

**Table 1.** Clinical and demographic summary of clear cell renal cell carcinoma cohorts.

| Variables | TCC ccRCC patients, no. (%) (n=105) | CPTAC-3 ccRCC patients, no. (%) (n=110) | TCGA-KIRC ccRCC patients, no. (%) (n=441) |
| --- | --- | --- | --- |
| **Sex** |  |  |  |
| Female | 35 (33.3) | 30 (27.3) | 153 (34.7) |
| Male | 70 (66.7) | 80 (72.7) | 288 (65.3) |
| **Race** |  |  |  |
| Asian Indian or Pakistani | 2 (1.9) | 1 (0.01) | 7 (1.6) |
| Black | 3 (2.9) | 1 (0.01) | 30 (6.8) |
| Other | 7 (6.7) | - | - |
| White | 93 (88.5) | 61 (55.5) | 397 (90.0) |
| Not Reported | - | 47 (42.7) | 7 (1.6) |
| **Tumor laterality** |  |  |  |
| Left | 62 (59.0) | NA | NA |
| Right | 43 (41.0) | NA | NA |
| **Surgery type** |  |  |  |
| Partial nephrectomy | 22 (21.0) | NA | NA |
| Radical nephrectomy | 65 (61.9) | NA | NA |
| Radical nephrectomy with thrombectomy | 18 (17.1) | NA | NA |
| **Fuhrman nuclear grade** |  |  |  |
| 1 | 0 | 7 (6.4) | 9 (2.0) |
| 2 | 30 (28.6) | 53 (48.2) | 182 (41.3) |
| 3 | 62 (59.0) | 41 (37.3) | 179 (40.6) |
| 4 | 13 (12.4) | 9 (8.2) | 68 (15.4) |
| Not Reported | - | - | 3 (0.7) |
| **pT** |  |  |  |
| T1 | 31 (29.5) | 52 (47.3) | 211 (47.8) |
| T2 | 3 (2.9) | 13 (11.8) | 58 (13.2) |
| T3+T4 | 70 (66.7) | 45 (40.9) | 172 (39.0) |
| Not Reported | 1 (1.0) | - | - |
| **pN** |  |  |  |
| N0 | 27 (25.7) | 16 (14.5) | 203 (46.0) |
| N1 | 5 (4.8) | 4 (3.6) | 13 (2.9) |
| NX | 73 (69.5) | 89 (81.9) | 225 (51.0) |
| **pM** |  |  |  |
| M0 | - | 34 (30.9) | 353 (80.0) |
| M1 | 23 (21.9) | 3 (2.7) | 73 (16.6) |
| MX | 82 (78.1) | - | 14 (3.2) |
| Not Reported | - | 73 (66.4) | 1 (0.2) |
| **Sarcomatoid Status** |  |  |  |
| No | 99 (94.3) | NA | NA |
| Yes | 6 (5.7) | NA | NA |
| **Vitality** |  |  |  |
| Alive | 82 (78.1) | 96 (87.3) | 118 (83.1) |
| Dead | 23 (21.9) | 14 (12.7) | 24 (16.9) |
| **Age at surgery (yr)** |  | **Age at diagnosis (yr)** | **Age at diagnosis (yr)** |
| Median (range) | 65 (36 - 87) | 60 (30 - 89) | NA |
| **Pathological tumor size (cm)** |  |  |  |
| Median (range) | 6.0 (1.3 - 17.5) | 6.4 (1.0 - 16.0) | NA |
| **% EGFR splice variant alpha** |  |  |  |
| Median (range) | 1.01 (0.00 - 41.33) | NA | NA |
| **BAP1 mutation** |  |  |  |
| Wildtype | 61 (58.1) | 93 (84.5) | 372 (84.4) |
| Alteration | 4 (3.8) | 17 (15.5) | 47 (10.6) |
| Not Reported | 40 (38.1) | - | 22 (5.0) |
| **KDM5C mutation** |  |  |  |
| Wildtype | 56 (53.3) | 91 (82.7) | 389 (88.2) |
| Alteration | 9 (8.6) | 19 (17.3) | 30 (6.8) |
| Not Reported | 40 (38.1) | - | 22 (5.0) |
| **MTOR mutation** |  |  |  |
| Wildtype | 62 (59.0) | 104 (94.5) | 392 (88.9) |
| Alteration | 3 (2.9) | 6 (5.5) | 27 (6.1) |
| Not Reported | 40 (38.1) | - | 22 (5.0) |
| **PBRM1 mutation** |  |  |  |
| Wildtype | 41 (39.0) | 66 (60.0) | 258 (58.5) |
| Alteration | 24 (22.9) | 44 (40.0) | 161 (36.5) |
| Not Reported | 40 (38.1) | - | 22 (5.0) |
| **PTEN mutation** |  |  |  |
| Wildtype | 60 (57.1) | 105 (95.5) | 399 (90.5) |
| Alteration | 5 (4.8) | 5 (4.5) | 20 (4.5) |
| Not Reported | 40 (38.1) | - | 22 (5.0) |
| **SETD2 mutation** |  |  |  |
| Wildtype | 55 (52.4) | 95 (86.4) | 360 (81.6) |
| Alteration | 10 (9.5) | 15 (13.6) | 59 (13.4) |
| Not Reported | 40 (38.1) | - | 22 (5.0) |
| **TP53 mutation** |  |  |  |
| Wildtype | 61 (58.1) | 104 (94.5) | 408 (92.5) |
| Alteration | 4 (3.8) | 6 (5.5) | 11 (2.5) |
| Not Reported | 40 (38.1) | - | 22 (5.0) |
| **VHL mutation** |  |  |  |
| Wildtype | 18 (17.1) | 28 (25.5) | 186 (42.2) |
| Alteration | 47 (44.8) | 82 (74.5) | 233 (52.8) |
| Not Reported | 40 (38.1) | - | 22 (5.0) |
| **Diabetes Status** |  |  |  |
| No | 76 (72.4) | NA | NA |
| Yes | 29 (27.6) | NA | NA |
| Not Reported | - | 110 | 441 |
| NA = no available; ccRCC = clear cell renal cell carcinoma; TCC = Total Cancer Care Protocol; CPTAC-3 = Clinical Proteomic Tumor Analysis Consortium 3; TCGA-KIRC = The Cancer Genome Atlas Kidney Renal Clear Cell Carcinoma |  |  |  |

To further investigate the trends identified in point-estimates of diversity from T-cell receptor (TCR) and B-cell immunoglobulin recoveries the Moffitt TCC patient bulk RNA-sequencing, we validated trends identified in the Moffitt TCC Cohort analysis with complementary analysis with the RNA-sequencing from the under CPTAC-3 Cohort (written consent had been obtained under CPTAC guidelines). Table 1 shows a summary of the clinical information obtained from individuals in the CPTAC-3 cohort. TCGA-KIRC RNAseq based TRA and TRB CRD3s were obtained from Thorsson et al (27, 28) based on the dbGAaP approved protocol number 6300. Relevant clinical and pathological outcomes, aligning with outcomes available in the Moffitt TCC Cohort, that are available in CPTAC-3 and TCGA-KIRC Cohorts are reported in Table 1.

### Recovery of immune receptor V(D)J recombination reads from bulk RNA-sequencing

Recovery of immune receptor V(D)J recombination reads was performed in two steps. First, RNAseq binary alignment map (BAM) files were searched, as a straight string search, for 10-mer nucleotide sequences representing the 3’ ends of every human V-gene and 5’ end of every human J-gene, for all seven immune receptors. Next, the resulting reads were aligned to reference V and J regions obtained from the International Immunogenetics Information System (IMGT). The quantitative parameters for the pairwise alignment were: (i) nucleotide match, + 5, (ii) mismatch, − 10, (iii) opening gap, − 10, and (iv) extending gap, − 10. The threshold for a V or J gene segment match was a score of ≥ 65. To ensure V and J read fidelity, only reads with a 20 nucleotide or greater match length for both V and J regions, and within the 20 nucleotides, a > 90% nucleotide match fidelity for both V and J regions were considered as matches. Additionally, and a productive CDR3 domain, defined as an in-frame junction without stop codons, was required for recombination read identification. Code for the method described above can be obtained at: https://github.com/bchobrut-USF/vdj under “Code Package A”. See also https://hub.docker.com/r/bchobrut/vdj for a container version of the code with a readme file.

### Generalized diversity index for patient quantifying CDR3 receptor diversity

The generalized diversity index (GDI) can be viewed as a continuous, non-increasing function over a range of values described by the parameter *q*, called order of diversity. This parameter allows a consideration of multiple scales of diversity simultaneously or in combination. GDI is often used in ecology (20) and was more recently introduced to quantify intratumor heterogeneity and evolution (29–31). Formally, GDI is calculated as:

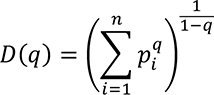

where *D*(*q*) is called the diversity index at the given *order of diversity q*, *n* is the number of unique CDR3 sequences recovered across the entire cohort, and *p*_!_ is the relative proportion of *i*-th CDR3 sequence. We typically evaluated the diversity score, *D*(*q*), for *q* between 0.01 and 100 numerically for each patient, for each of the receptor and immunoglobulin types individually, as well as in biologically meaningful groups (TRA+TRB together, TRG+TRD together, and IGH+IGK+IGL together). Varying the value of *q* represents interpolating between richness and evenness: richness is weighted more at low values of q, and evenness is weighted more at higher values of q. High-q GDI scales inversely with dominance or clonality.

Point-estimates derived from patient’s GDI, D(q), can be used for easy of comparison of sequence distributions across patients and cohorts. These point-estimates of interest include (i) low-q diversity (*D*(0.01)), which describes the epitope richness of the patients, (ii) high-q diversity (*D*(100)), which describes dominance of the “main driving” epitope, and (iii) ΔD, which measures the difference between low-q and high-q diversity (*D*(0.01) − *D*(100)). Furthermore, when visualizing the continuum of diversity measures *D(q)* with *q* in log-scaling, the continuum of diversity measures appears to have an inflection point, corresponding to a scale of diversity where small changes in the key parameter *q* can have large impact: the higher this value, the more even we expect a distribution to be, as the inflection point tends to infinity for perfectly even distributions (corresponding to *n* sequences all at frequency 1/*n*). Thus, two additional point-estimates of interest that we used are (iv) the value *q* at which an inflection point occurs (IP), and (v) the slope at the inflection point (denoted as IP slope). All code for calculating CDR3 diversity and its summary metrics has been implemented in Julia (version 1.4.0) and documented in the publicly available package OncoDiversity.jl.

To determine the impact of all five point-estimates of diversity, we ran a correlation analysis and determined that we could reduce our five point-estimates of diversity down to three metrics for comparison across receptor groups and patients. The Spearman correlation coefficients were calculated between point-estimate metrics and comparisons between low-q diversity, ΔD diversity, and the inflection point slope all had significant and very strong Spearman correlation coefficients of 0.98 or greater, so moving forward, we just focused on one of those metrics as a measure of species richness diversity (Supplemental Fig. S2). There was not a strong correlation between high-q diversity and the inflection point q metrics and either metric compared to any of the three species richness diversity metrics (low-q diversity, ΔD diversity, and inflection point slope), so we continue to look at the high-q diversity and inflection point q separately and in additional to the single species richness measure.

### Assessment of clinical and survival associations with CDR3 features

Clinical associations were evaluated for recoveries in TRA, TRB, TRG, TRD, IGH, IGK, and IGL separately as well as in combinations of TRA+TRB, TRG+TRD, and IGH+IGK+IGL. After point estimates of diversity were calculated for each patient and each receptor subtype/combination, the clinical parameter values were assessed to identify if CDR3 receptor diversity could discriminated ccRCC patients based on relevant clinical and pathological outcomes, as well as the percentage of tumor with EGFR spice variant alpha. Largest diameter size and age were the only two continuous variables, which were evaluated by dividing the cohort into above and below the median of the diversity and compared with unpaired t-tests. All other categorial data types were divided by categories and the diversity metric was compared across the categories using unpaired t-test for 2 categories and ANOVA for 3 or more categories.

Survival correlations for the above combinations were performed by separating the cohort into above and below the median based on point-estimates of the generalized diversity index. In addition, the maximally selected rank statistical analysis was performed to estimate an optimal cut point in the quantitative point estimates as a binary classification rule regarding overall survival time (32). The Kaplan-Meier (KM) curve method was used to calculate survival probability and log-rank test was used to compare the above (high diversity) and below (low diversity) groups. Graph-Pad Prism software (version 8) and R version 3.6.1 were used for computing statistical comparisons and outputting figures.

### xCell Scores

From bulk RNA-sequencing for each patient, xCell scores were calculated for various T-cell and B-cell subtypes as well as the Immune Score, Stroma Score, and Microenvironment Score. Then for each xCell score calculated a Spearman correlation was calculated for the total and unique recoveries identified for TRA receptor, IGL receptor, and aggregate combination (TRs+IGs).

Patients in the Moffitt TCC cohort had previously undergone bulk RNA sequencing of macro-dissected tumor samples using the TruSeq RNA Exome kit (Illumina) for 50 million 100– base pair paired-end reads. RNA sequence reads were aligned to the human reference genome in a splice-aware fashion using Spliced Transcripts Alignment to a Reference (STAR) (26), allowing for accurate alignments of sequences across introns. Aligned sequences were assigned to exons using the HTseq package (33) to generate initial counts by region. Normalization, expression modeling, and difference testing were performed using DESeq2 (34). For the CPTAC cohorts, detailed methodology regarding RNA sequencing can be found at its source web page(25).

RNA sequencing data was analyzed for cell-type enrichment using the xCell bioinformatic pipeline (25). xCell uses a compendium of validated gene expression signatures for 64 individual cell-types derived from thousands of expression profiles. Single-sample gene set enrichment analysis scores were adjusted for spillover compensation to generate an adjusted enrichment score for each cell type within the specimen, which is referred to as the xCell score. xCell scores were generated for each of the 64 cell-types for each ccRCC tumor specimen.

## RESULTS

### General diversity index (GDI) quantifies tumor infiltrating lymphocyte receptor subtype diversity in the TCC cohort

For each patient, we measured individual receptor CDR3 diversities across the 7 human adaptive immune receptor genes (TRA, TRB, TRG, TRD, IGH, IGK, IGL), as well as common combinations of these receptor subtypes (TRA+TRB, TRG+TRD, IGH+IGK+IGL, along with all 7 together, denoted at TRs+IGs). In the TCC cohort (n = 105), CDR3 sequences were recovered from bulk RNA-sequencing of patient tumor tissue. GDI was then calculated for each subtype and group of subtypes (the landscape of recoveries across common groups are shown in Fig. 1A-B and Supplemental Fig. S3 and distribution of individual recoveries in Supplemental Fig. S4). The Moffitt TCC cohort of ccRCC patients represented a cohort of clinically high-risk and advanced patients. Over two-thirds of the cohort contains patients with pathologic stages 3 or 4, including 6% of patients with highly aggressive sarcomatoid histology (Table 1). To quantify the GDI, we generated a continuum of diversity measures D(q) for each patient across values of the order of diversity, q. Then, we were able to compare clinical variables at specific point estimates of the continuum of diversity measures, as shown in Fig. 1C. We compared immune receptor subtype diversity across patients, and found that the point estimates ΔD diversity, high-q diversity, and inflection point (IP) of the GDI curve (see Methods) were unique summary metrics. The value of ΔD summarizes richness (total number of unique sequences) of receptor subtypes in a patient sample. High-q diversity focuses on the dominance (frequency of largest sequence) of a receptor subtype and de-emphasizes a rare receptor subtype. IP is a measure of receptor subtype evenness; with high IP values indicating an overall more level distribution, largely independent of receptor subtype richness (29–31).

**Figure 1:**
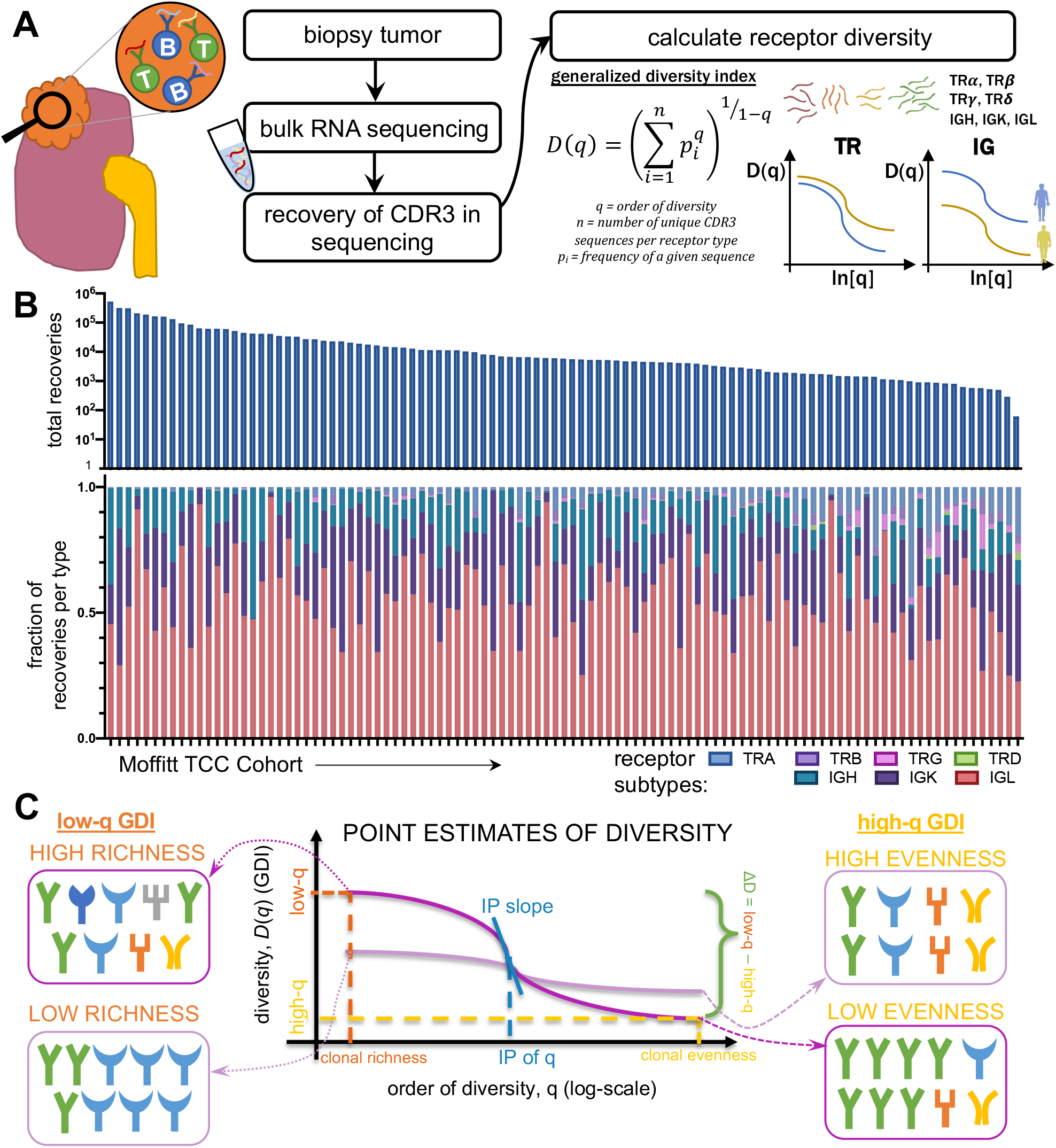
Tumor infiltrating lymphocyte receptor diversity as a marker in clear cell renal cell carcinoma. Overall workflow schematic of calculating tumor infiltrating lymphocyte (TIL) diversity across a cohort of patients. Patient tumors undergo bulk RNA sequencing and then CDR3 sequences from TCR and BCR receptors are recovered. Then for each patient, CDR3 sequences are segregated by receptor class (TRA, TRB, TRG, TRD, IGH, IGK, and IGL) and then patient frequencies across the CDR3 landscape per receptor are calculated and used to quantify the individual patient’s receptor diversity using the generalized diversity index from ecology. (B) Receptor recovery distributions across the seven major receptors types in the Moffitt cohort consented under the Total Cancer Care (TCC) protocol. (C) Patient diversity curves can be distilled down to five point-estimates of diversity: low q (q=0.02), high q (q=100), ΔD (D(0.01)–D(100)), inflection point (IP), and inflection point slope.

### Immune receptor subtype richness is associated with important pathological features in the TCC cohort

Across individual receptor subtypes, TRA and IGL receptor diversity consistently showed increased richness (in Fig. 2 exemplified with ΔD diversity comparisons) in tumors with larger diameters, higher grade, sarcomatoid status, and tumors from the left side. TRA receptor diversity split the Moffitt TCC cohort at the median of ΔD diversity. Of these, patients with ΔD values below the cohort median had a mean largest diameter size of 6.1 cm, compared to those with above the median who had a mean largest diameter size of 7.6 cm (Fig. 2A i, p-value: 0.0287). This same trend was reflected in IGL receptor diversity with the high ΔD diversity group (Fig. 2B i, p-value: 0.0195).

**Figure 2:**
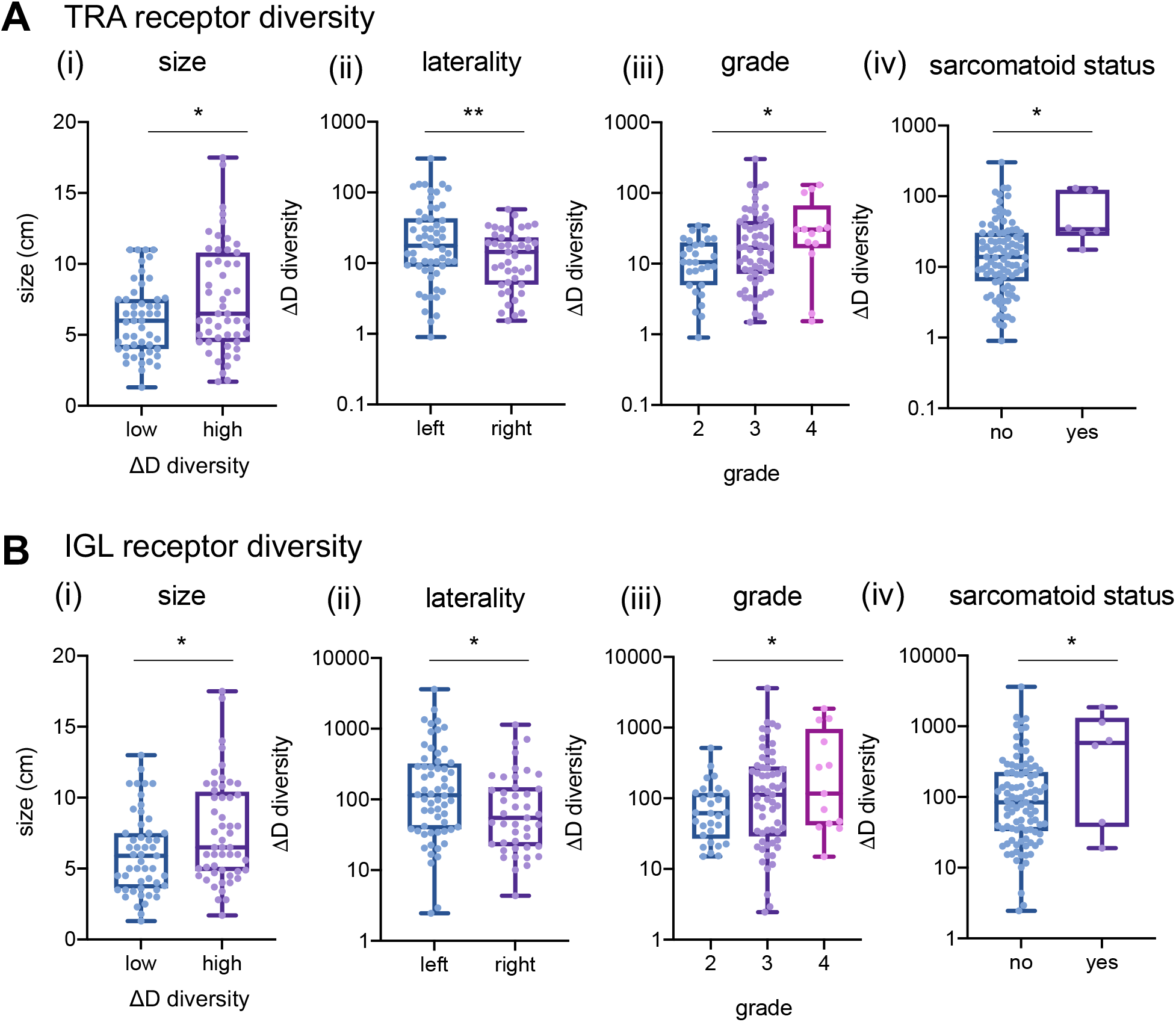
Patients with tumors that are larger in diameter, higher grade, left laterality and sarcomatoid status have increased diversity in TRA and IGL receptors. (A) TRA receptor CDR3 sequence ΔD diversity across the Moffitt TCC Cohort have increased diversity in (i) larger diameter tumors (low diversity had mean diameter of 6.1 cm and high diversity had a mean diameter for 7.6cm; p-value: 0.0287), (ii) left laterality tumors (score of 37.55 vs 16.39; p-value: 0.0097), (iii) with higher grade tumors (mean score from grade 2 was 12.96, mean score from grade 3 was 33.09, and mean score from grade 4 was 42.62; p-value: 0.0227), and (v) in patients with sarcomatoid status (sarcomatoid status evaluated as yes (at least 5%) or no, in TRA no had a mean ΔD diversity score of 26.47 vs yes with a mean score of 61.32; p-value 0.0430). (B) IGL receptor CDR3 sequence ΔD diversity showed the same trends as TRA receptor CDR3 sequence diversity for (i) size, (ii) laterality (score of 331.1 vs 141.3; p-value: 0.0445), (iii) grade (mean score from grade 2 was 93.99, mean score from grade 3 was 281.4, and mean score from grade 4 was 465.5; p-value: 0.0459), and (iv) sarcomatoid status (no had a mean score of 223.9 vs yes with a mean score of 704.4; p-value: 0.0152). Unpaired t-tests were used to compare two group data and ANOVA was used to compare grade, three group data. p-value significance represented by * < 0.05, ** < 0.01, *** < 0.001

TRA receptor ΔD diversity in CDR3 amino acid sequences recovered from tumors with left laterality had an average diversity score 2.3-fold higher compared to those with right laterality tumors (Fig. 2A ii, p-value: 0.0097), which was also reflected in IGL receptor ΔD-diversity (Fig. 2B ii, p-value: 0.0445). In addition, patients with high tumor grade had increased TRA receptor ΔD diversity (Fig. 2A iii, p-value: 0.0227), which was also demonstrated in IGL receptor ΔD diversity (Fig. 2B iii, p-value: 0.0459).

Overall, Moffitt TCC cohort tumors that were identified with sarcomatoid histology had increased overall lymphocyte receptor diversity compared to those individuals who did not have sarcomatoid histology (demonstrated in Fig. 2A & B iv; p-value: 0.0430 in TRA and p-value: 0.0152 in IGL). This trend for increased diversity in sarcomatoid carcinoma tumors was statistically significant in all combinations except for TRG receptor diversity, which was one of the rarest CDR3 receptor subtypes recovered.

Our observations of increased lymphocyte receptor richness in larger diameter tumors, higher grade tumors, left laterality tumors, and sarcomatoid carcinomas were also discovered in other receptor subtypes, as well as in the combinations (Supplemental Fig. S5 shows size, laterality, grade, and sarcomatoid status across all combinations of receptors, Supplemental Fig. S6 shows the Shannon index of TRA and IGL across size, laterality, grade, and sarcomatoid status and **Supplemental File 1** contains statistics for all comparison combinations across all clinical features).

### Independent validation of GDI metrics

To validate the findings of increased diversity with poor pathological features we first calculated xCell scores (35) for patients in the Moffitt TCC cohort to confirm that the detected recoveries came from tumor-infiltrating lymphocytes. TRA receptor total and unique recoveries had the highest Spearman correlation scores with T-cell subtype xCell score and were less correlated with the B-cell subtype xCell score (Fig. 3A and full correlation analysis of all xCell scores with total and unique recoveries of TRA is shown in Supplemental Fig. S7, with total and unique recoveries of IGL in shown in Supplemental Fig. S8, and with total and unique recoveries of all CDR3s recovered is shown in Supplemental Fig. S9). The strongest correlation associated with TRA receptor was with CD8+ Tcm (r=0.819 with total recoveries; r=0.714 with unique recoveries) and CD8+ T-cells (r=0.751 with total recoveries; r=0.695 with unique recoveries), while the weakest correlation was with CD4+ Tcm (r=0.177 with total recoveries; r=0.185 with unique recoveries) and CD4+ naïve T-cells (r=0.243 with total recoveries; r=0.097 with unique recoveries). These correlations also held true for IGL receptor recoveries and total (TRs+IGs) receptor recoveries, but the correlations were moderate in strength (most Spearman correlation coefficients between 0.2-0.5) compared to the Spearman correlation coefficient associated with TRA (most correlation coefficients between 0.3-0.7). Furthermore, the Immune Score strongly correlated positively with total and unique recoveries, compared to the Microenvironment Score and Stroma Score, which both showed weaker Spearman correlation coefficients.

**Figure 3.**
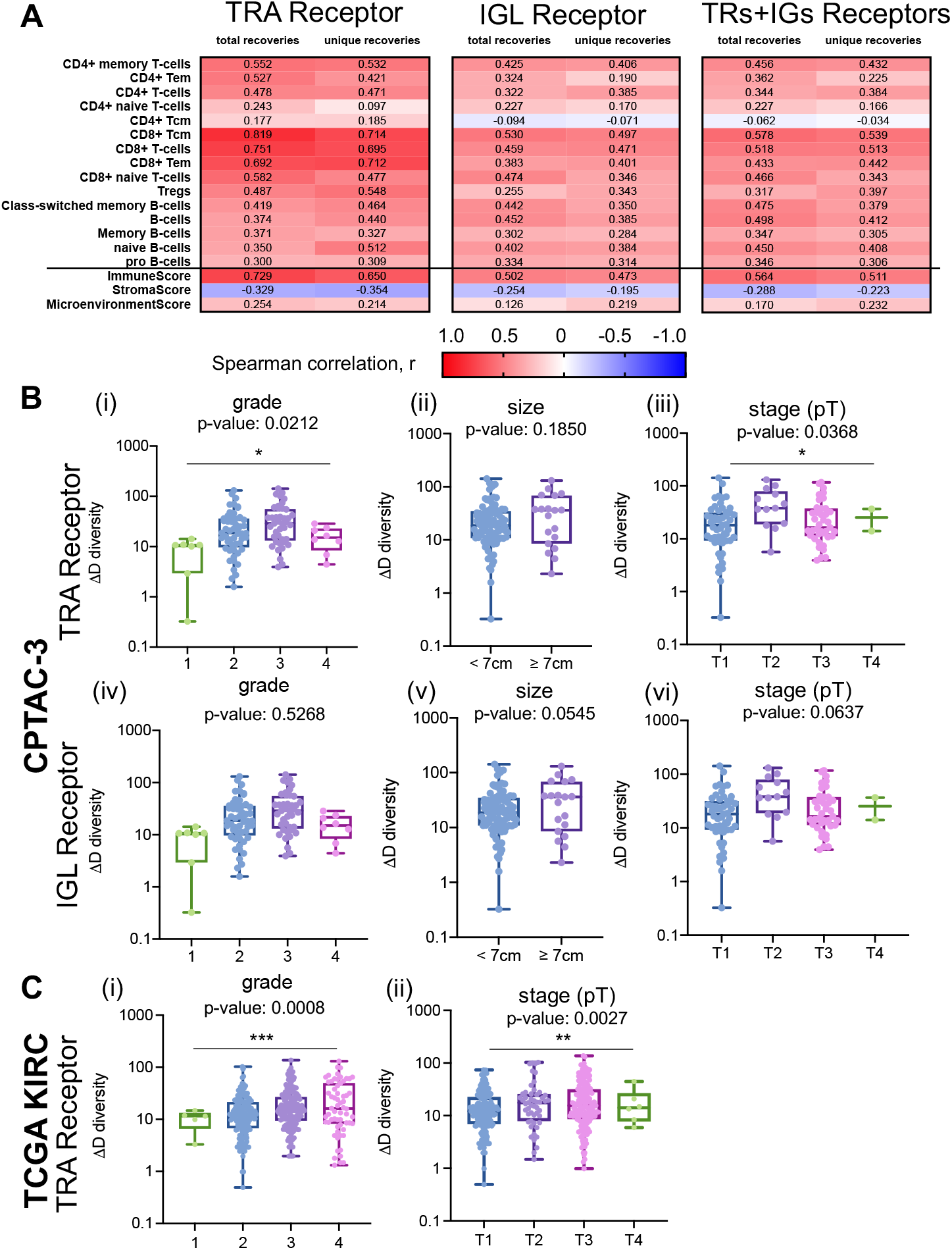
CDR3 sequence diversity trends were validated using xCell scores and a secondary RCC CPTAC3 cohort. (A) Spearman correlation coefficient, r, was calculated between the total and unique number of TRA, IGL, and total (TRs+IGs) recoveries and the xCell scores for various T-cell and B-cell subtypes, Immune Score, Stroma Score, and Microenvironment Score. (B) Grade, largest diameter size, and stage (pT) trends in TRA receptor and IGL receptor ΔD diversity in the CPTAC-3 cohort. For TRA recoveries, patients (n= 108) had increased ΔD diversity in (i) higher grade (mean score from grade 1 was 8.558, mean score from grade 2 was 27.94, mean score from grade 3 was 38.60, and mean score from grade 4 was 15.92; p-value: 0.0212), (ii) a higher mean ΔD diversity score in tumors with diameters greater than or equal to 7cm (mean score in diameters ≥ 7cm was 37.91, mean score in diameters < 7cm was 28.12; p-value: 0.1850), and (iii) in more advanced pT stage (mean score from T1 was 25.01, mean score from T2 was 51.07, mean score from T3 was 29.24, mean score from T4 was 5.32 p-value: 0.0368). Similarly, for IGL recoveries, patients had increased ΔD diversity in (iv) higher grade (mean score from grade 1 was 50.14, mean score from grade 2 was 160.6, mean score from grade 3 was 156.9, and mean score from grade 4 was 84.79; p-value: 0.5268), (v) a higher mean ΔD diversity score in tumors with diameters greater than or equal to 7cm (mean score in diameters ≥ 7cm was 235.6, mean score in diameters < 7cm was 126.6; p-value: 0.0545), and (vi) in more advanced pT stage (mean score from T1 was 147.0, mean score from T2 was 291.1, mean score from T3 was 97.58, mean score from T4 was 120.5; p-value: 0.0637). (C) Grade and stage (pT) trends in TRA receptor ΔD diversity in the TCGA KIRC cohort. Patients (n=390) had increased ΔD diversity in (i) higher grade (mean score from grade 1 was 10.28, mean score from grade 2 was 16.62, mean score from grade 3 was 22.49, and mean score from grade 4 was 28.32; p-value: 0.0008), and (ii) in more advanced pT stage (mean score from T1 was 16.64, mean score from T2 was 24.25, mean score from T3 was 24.78, mean score from T4 was 17.90; p-value: 0.0027). Unpaired t-tests were used to compare two group data and ANOVA was used to compare grade, three group data. p-value significance represented by * < 0.05, ** < 0.01, *** < 0.001.

Once we confirmed that the diversity scores, we measured were attributable to the immune cell infiltration in the tumor with the xCell scores, we sought to independently validate our findings with a replicative study, using the ccRCC CPTAC-3 cohort (n=110). The CDR3 recovery landscape of CPTAC-3 and Moffitt TCC differed slightly. CPTAC-3 had more recoveries from T-cells; B-cells accounted for an average of only 78.96%. However, we did not identify any significant trend of differences between the proportion of T-cell and B-cell recovered based on stage (CPTAC-3 recovery landscape is shown in Supplemental Fig. S10 and fraction of B-cell and T-cell receptors recovered per patient grouped by stage is detailed in Supplemental Fig. S11). Laterality and sarcomatoid status could not be evaluated, as these are not available in CPTAC-3.

We confirmed the observation that higher grade and larger size of tumors are associated with increases in TRA and IGL receptor ΔD diversity in the CPTAC-3 cohort (Fig. 3B compared to Supplemental Fig. S12, and **Supplemental File 2** contains statistics for all comparisons across all clinical features)—which independently validated that rich, but rather uneven receptor subtype distribution is associated with larger and high-grade tumors. TRA receptor ΔD diversity was significantly different between low and high grade (Fig. 3B i, p-value: 0.0212) and pT stages (Fig. 3B iii, p-value: 0.0368). In both TRA and IGL receptor distributions, the ΔD diversity score showed that large tumors (described pathologically as tumors with the largest diameter 7 cm or greater) have increased receptor subtype diversity compared to smaller tumors (those with largest diameters below 7 cm). Of note, CPTAC-3 has significantly different distribution of tumor sizes, with many small tumors (largest diameter < 7cm), compared to the Moffitt TCC cohort (Fig. 3B ii and v, Supplemental Fig. S13 compares the Moffitt TCC and CPTAC-3 Cohorts based on distribution of tumor sizes).

Furthermore, the differences in grade and stage (size data not available) could be replicated in the TRA recoveries from previously obtained TCR CDR3s(27, 28) from The Cancer Genome Atlas Kidney Renal Clear Cell Carcinoma (TCGA-KIRC) Cohort. As demonstrated in the Moffitt TCC Cohort (Fig. 2A iii and Supplemental Fig. S12A iii) and CPTAC-3 Cohort (Fig. 3B i, iii), TCGA KIRC Cohort patients with higher grade (Fig. 3C i, p-value: 0.0008) and higher pT stage (Fig. 3C ii, p-value: 0.0027) had significantly higher TRA receptor ΔD diversity.

In addition to tumor sample differences, CPTAC-3 Cohort had a subset of 75 patients with matched normal tissue samples (matched normal tissue recovery landscape described in Supplemental Fig. S14). Lymphocyte receptor richness was increased in tumor samples compared to normal tissue, which was observed across all receptor subtypes and combinations (Fig. 4A, Supplemental Fig. S15, and **Supplemental File 2**). In the TRs+IGs combination, tumor tissues had on average, at least 2.6-fold increase in richness of CDR3 sequences recovered, compared to the matched patient’s normal tissue (mean score of 144.0 in normal tissue compared to a mean score of 377.1 in matched tumor). Furthermore, sequence dominance (measured by low values of high-q diversity) was decreased in all receptor subtypes except for the IGL receptor and IGH+IGK+IGL receptor combination (Fig. 4B, Supplemental Fig. S15, and **Supplemental File 2**). In the TRs+IGs combination, the tumor tissue had on average, at least 1.3-fold increase in high-q diversity compared to normal tissue, which indicates that the most abundance CDR3 sequence in the sample was about 2% lower, thus less dominant, in the tumor (mean score of 14.27 in the normal tissue compared to a mean score of 19.17 in the tumor tissue). Analyzing the inflection point q-metric of evenness in TRs+IGs combinations in the normal-tumor matched patients showed that normal samples have an almost 10% mean increase evenness compared to their matched tumor samples (Fig. 4C; p-value 0.0857). This data supported the hypothesis that normal tissues are expected to show very even CDR3 sequence distributions, and that better outcomes are to be expected in tumors that appear more normal in this context.

**Figure 4:**
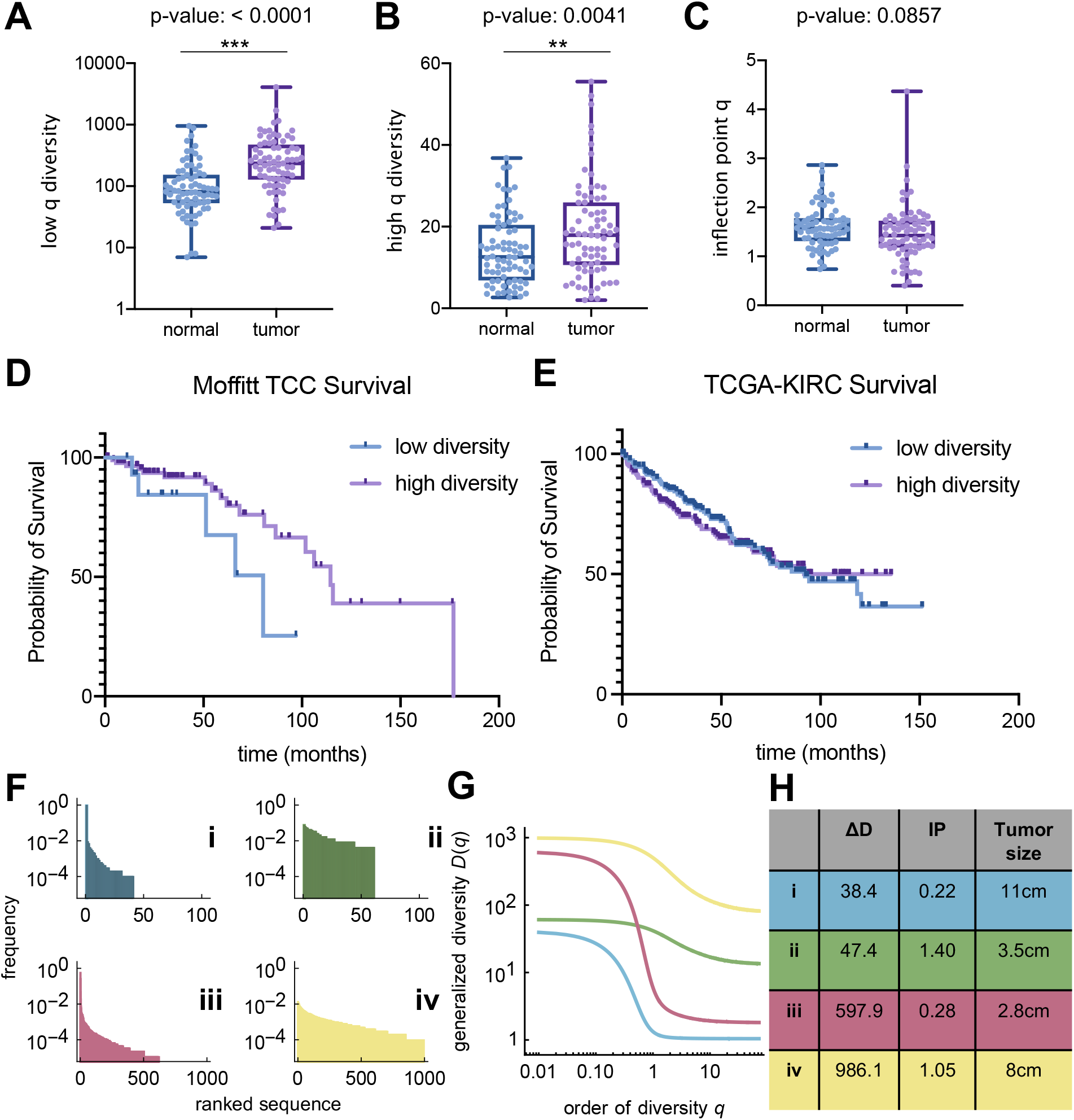
Novel measures of diversity and overall survival. Tumor samples have increased (A) TRs+IGs (all receptor combination) species richness and (B) evenness of CDR3 receptor sequences compared to patient-matched normal tissue. However, tumor samples have (C) reduced TRs+IGs inflection point q diversity compared to normal tissue (mean of normal 1.602 vs mean of tumor 1.465; p-value 0.0857). (D) Individuals in the Moffitt TCC Cohort with larger TRA distribution inflection point (IP, see Methods) had significantly improved overall survival (hazard ratio: 0.526, Cox p-value: 0.036) with a median survival of 115 months compared to those with lower IP. (E) TCGA-KIRC overall survival supports the trend of lower IP has reduced median survival (92.13 months compared to undefined in high IP group; log-rank p-value: 0.6104). (F,G,H) Four example patients of the Moffitt TCC cohort with fundamentally different characteristics. While ΔD assesses mainly sequence richness, the inflection IP, clearly visible in (G), is a robust measure of evenness: high IP-distributions are more even.

### Immune receptor subtype evenness, measured by IP, is associated with survival

We first analyzed associations between immune receptors and overall survival in the Moffitt TCC Cohort. Each of our chosen diversity metrics (**Methods**) was compared for each of the 7 immune receptor types. We used the maximally selected rank statistics (maxstat) approach (Methods), with a cut-point that yielded a maximal survival difference, together with a multivariate Cox regression analysis. We found larger TRA sequence distribution IP, which measures distribution evenness, was significantly associated with longer overall survival (Fig. 4D). The IP value of the cut-point in the cohort that yielded a maximal survival difference was 0.826. Using this optimal cut-point resulted in a hazard ratio of 0.526 (Log-rank p-value: 0.049, Cox p-value: 0.036) with the low IP group (IP <0.826, n=15) having a median overall survival of 80 months, and the high diversity group (IP >0.826, n=88) having a median overall survival of 115 months. This trend could not be confirmed with the CPTAC-3 cohort due to the lack of survival information, i.e., overall survival data information was censored for 85 of the 98 CPTAC-3 cases (Supplemental Fig. S16). This trend was supported (although not statistically significantly) with the TCGA-KIRC cohort with individual with IP above the median produced a low IP group (IP<8.26, n=192) having a median overall survival of 92.1 months, and the high IP group (IP>8.26, n=197) having an undefined median overall survival due to survival fraction not falling below 50% in this group, which is likely due to this cohort also being comprised of lower grade and stage tumors compared to the Moffitt TCC cohort (Fig. 4E; log-rank p-value: 0.6104).

To further illustrate the utility of our diversity metrics for receptor subtype heterogeneity estimation, we show in Fig. 4F-H four Moffitt TCC cohort examples of characteristic differences in richness versus evenness space. In both examples of low evenness, dominance (low high-q diversity) is high (Fig. 4G). Interestingly, the survival difference unveiled by IP distribution evenness comparison does not necessarily coincide with tumor size, as size rather correlated with richness (ΔD, Fig. 4H). Taken together, these findings highlight the ability of these metrics to assess immune sequence distribution heterogeneity via GDI-derived point estimates.

### Demographic and aberrant splicing differences in tumor-infiltrating lymphocyte receptor diversity

In addition to capturing differences in tumor pathology and survival, the GDI was also able to discriminate patients based on demographic differences. B-cell receptor high-q diversity is an estimate of dominance by the most abundant sequence: the lower this value, the more dominant the most abundant sequence. High-q diversity demonstrated differences in immune infiltration of white versus non-white ccRCC patients in the TCC cohort (Supplemental Fig. S17). IGH and IGL receptor high-q diversity alone, as well as the IGH+IGK+IGL and total recovery (TRs+IGs) showed at least a 2.1-fold increase in non-white patients compared to white patients (Supplemental Fig. S17A-B; p-values < 0.01). This trend was also observed in the IGK receptor high-q diversity, but only with a 1.6-fold increase in non-white individuals (Supplemental Fig. S17C; p-value 0.0561).

Furthermore, diversity metrics from T-cell receptors TRG and TRD discriminate patients based on gender differences in the TCC cohort. TRG and TRD receptor inflection point q was at least 1.7-fold higher in female patients compared to male patients (TRG: score of 2.503 in females vs. a score of 1.442 in males, p-value: 0.0033; TRD: score of 2.883 in females vs. a score of 1.454 in males, p-value: 0.0003; Supplemental Fig. S18). Moreover, when high-q diversity (dominance of the most abundant sequence) was compared between female and male patients with respect to TRG and TRD diversity, in both the Moffitt TCC and CPTAC-3 cohorts, female patients had increased high-q diversity compared to male patients (Supplemental Fig. S19).

We evaluated association of GDI with a recently described aberrant EGFR splice variant in ccRCC (36). The Moffitt TCC Cohort of patients were profiled for the presence of this EGFR variant (reported as % EGFR variant), and we found a positive correlation between the percentage of tumors expressing the EGFR variant and the evenness (inflection point q) scores from B-cell receptors (Supplemental Fig. S20). This correlation was most significant in IGL receptor diversity, with a Spearman correlation coefficient of r=0.3430 (p-value: 0.004, B-cell receptor IP vs. EGFR variant correlation analysis is demonstrated in Supplemental Fig. S20 and T-cell receptor IP vs. EGFR variant correlation analysis is demonstrated in Supplemental Fig. S21), indicating more even (high IP) distributions have higher proportion of cells with that EGFR variant. This positive correlation between IP and percentage of EGFR variant was not as strong nor significant in the T-cell receptors (Supplemental Fig. S21).

To determine if pre-existing host inflammatory environment contributed to differences in patient CDR3 diversity, we identified patients in the Moffitt TCC cohort who were diagnosed with diabetes and investigate if there were any associations with the diversity metrics and diabetes status. We found that none of the 11 receptor combinations explored had a significant difference in any of the metrics of CDR3 diversity (individual comparisons are reported in **Supplemental File 1** and TRA, IGL, and TRs+IGs ΔD and IP comparisons are demonstrated in Supplemental Fig. S22).

Finally, we evaluated associations of GDI across the mutational landscape across all three cohorts. For each of the cohorts, we had mutational status on a subset of patients for common driver mutations in ccRCC including: BAP1, SETD2, KDM5C, MTOR, PBRM1, PTEN, TP53, and VHL. Across all three cohorts, the number of mutations in this shortlist of driver mutations was negatively correlated with richness (Fig. 5A). Interestingly, the Moffitt TCC cohort, generally considered the more aggressive cohort, had the fewest number of associations between diversity metrics and mutation status, while the CPTAC-3 and TCGA-KIRC cohorts had more significant associations (detailed for CPTAC-3 in **Supplemental File 2** and TCGA-KIRC in **Supplemental File 3**). Furthermore, B-cell receptors (IGH, IGK, IGL) had more significant associations with mutational status than T-cell receptors (TRA, TRB, TRG, TRD; compared between the Moffitt TCC cohort described in **Supplemental File 1** and CPTAC-3 cohort in **Supplemental File 2**). Specifically, with IGL recoveries in the CPTAC-3 cohort in patients with mutations in KDM5C, PBRM1, and VHL all had reduced richness (Fig. 5B i-iii) and mutation in PTEN was associated with increased evenness (Fig. 5B iv). These trends were confirmed in the total (TRs+IGs) recoveries of the CPTAC-3 cohort (Supplemental Fig. S23A) and the trends supported in by the Moffitt TCC cohort in direction, however they were not statistically significant (Fig. 5C for IGL trends and Supplemental Fig. S23B for TRs+IGs trends). In T-cell recoveries, both the CPTAC-3 and TCGA-KIRC cohorts showed reduced richness was associated with a mutation in PBRM1 (Fig. 5D), however this trend was not observed in the Moffitt TCC cohort.

**Figure 5.**
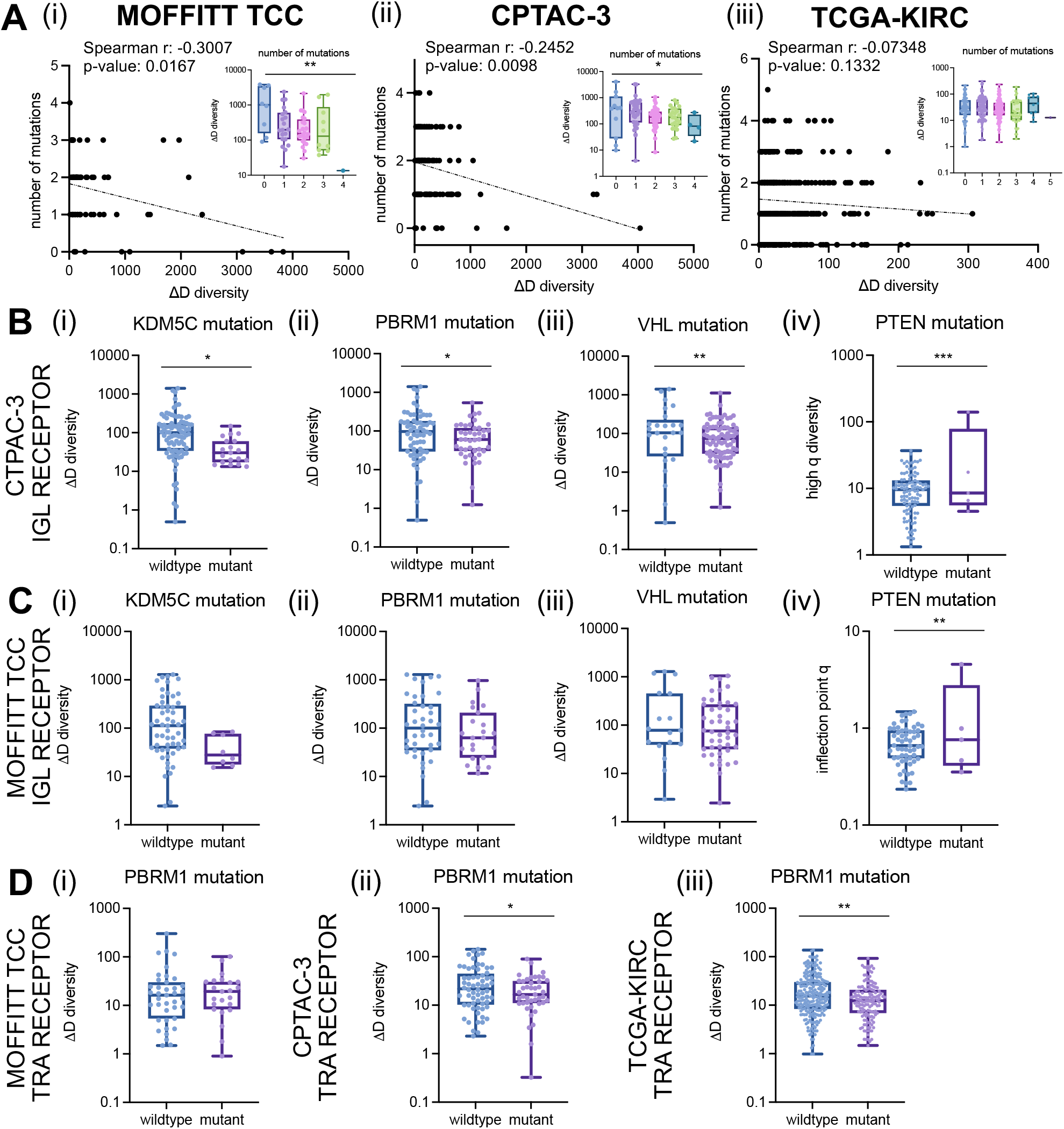
Associations of diversity metrics with mutational landscape in clear cell renal cell carcinoma. (A) Number of mutations were negatively correlated with CDR3 recovery richness. Spearman correlation coefficients between number of mutations and ΔD diversity were calculated for the (i) Moffitt TCC total (TRs+IGs) recoveries (Spearman r: -0.3007, p-value: 0.0167; ANOVA p-value: 0.0025); CPTAC-3 total recoveries (Spearman r: -0.2452, p-value: 0.0098; ANOVA p-value: 0.0213), and (iii) TCGA-KIRC total available (TRA+TRB) recoveries (Spearman r: -0.07348, p-value: 0.1332; ANOVA p-value: 0.3905). (B) IGL recoveries had reduced richness in CPTAC-3 patients with (i) KDM5C mutations (mean score of wildtype was 167.9 and mutant was 44.41; p-value: 0.0298), (ii) PBRM1 mutations (mean score of wildtype was 182.2 and mutant was 92.06; p-value: 0.0421), and (iii) VHL mutations (mean score of wildtype was 247.7 and mutant was 115.0; p-value: 0.0094) and increased evenness in patients with (iv) PTEN mutations (mean score of wildtype was 10.61 and mutant was 35.35; p-value: 0.0001). (C) IGL recoveries had reduced richness in Moffitt TCC patients with (i) KDM5C mutations (mean score of wildtype was 248.1 and mutant was 42.96; p-value: 0.0906), (ii) PBRM1 mutations (mean score of wildtype was 259.8 and mutant was 156.4; p-value: 0.2200), and (iii) VHL mutations (mean score of wildtype was 307.7 and mutant was 190.4; p-value: 0.1993) and increased evenness in patients with (iv) PTEN mutations (mean score of wildtype was 0.7323 and mutant was 1.427; p-value: 0.0083). (D) TRA receptor richness was (i) not different in Moffitt TCC patients with PBRM1 mutations (mean score of wildtype was 32.03 and mutant was 24.59; p-value: 0.5397), (ii) decreased in CPTAC-3 patients with PBRM1 mutations (mean score of wildtype was 33.63 and mutant was 22.53; p-value: 0.0470), and (iii) decreased in TCGA-KIRC patients with PBRM1 mutations (mean score of wildtype was 23.84 and mutant was 17.06; p-value: 0.0032). Unpaired t-tests were used to compare two group data and ANOVA was used to compare grade, three group data. p-value significance represented by * < 0.05, ** < 0.01, *** < 0.001.

## DISCUSSION

Here, we demonstrate how a generalized diversity index (GDI), often used in ecology and evolution (31,37,38), can be applied to quantitatively characterize tumor infiltrating lymphocyte receptor subtype diversity in ccRCC. We identified point estimates of this index that are associated with important differences in patient demographics, tumor pathology, and survival. These metrics can help objectively characterize host differences in immune receptor subtypes in ccRCC patients. These novel objective metrics can provide insight into underlying tumor and host immune relationships by defining differences within and across patients. We used bulk sequencing data from ccRCC tumors to better understand these differences in patient immune receptor subtypes and these metrics can be replicated in other similar cohorts with available sequencing data. These host diversity metrics could be especially helpful in elucidating the ideal tumor microenvironment for response to immunotherapy agents in patients with metastatic ccRCC (39).

In contrast to more numerous recombination read recovery, the recovery of adaptive immune receptor recombination reads from RNAseq files is obtained via PCR amplification of adaptive immune receptors. This procedure is also called the immune repertoire approach (40). Our work here strongly indicates the specific value of the TRA and IGL GDI, discussed in more detail below. Thus, it has made sense to apply the immune repertoire approach to increase the number of recombination reads for all adaptive immune receptors. We expect that other immune receptor genes may have prognostic value, with more recombination reads to evaluate. Ιmmune repertoire approaches, particularly when applied to cancer samples, often result in a majority of reads that represent relatively few clonotypes. In addition, human aging substantially reduces clonotype diversity (41, 42), particularly Figs. 2C and 2D therein. Thus “sampling of the repertoire”, by mining genomics files over large patient databases, can generate conclusions regarding clonotype associations with clinical features. More recent preparations of RNAseq files, including those we use here, have become much more robust over the last several years, both in terms of read quantity and lengths (28). Recovery of adaptive immune receptors from those files also has become more robust. In summary, our results from adaptive immune receptor read recoveries from the RNAseq files are informative. Further studies using the immune repertoire approach, representing a more comprehensive clonotype collection, should be applied in the future.

Our findings suggest that individuals with more advanced disease have increased richness in tumor recovered CDR3 sequences. In the Moffitt TCC Cohort, we detected statistically significant differences in TRA and IGL diversity with increased richness in tumors with larger diameter and higher grade. Furthermore, we identified tumors with sarcomatoid carcinoma pathology that represent a rare and aggressive histology and showed significant increases in different diversity metrics and receptor subtypes, in particular increased CDR3 sequence richness. The immune receptor profiles of these tumors are particularly interesting since sarcomatoid histology has also been associated with very favorable response to checkpoint inhibitors. We postulate that a similar immune receptor profile in other patient tumors may portend a favorable response to checkpoint inhibition. Also, we found a significant increase in richness in left sided tumors. This difference may explain some of the host related factors associated with left-sided tumors that have a poorer clinical outcome than right-sided tumors (43). Many of these associations were able to be validated in similar comparisons with the CPTAC-3 cohort studies.

In the TCC cohort, B-cell receptors with IGL had the most recoveries and TRA had the most T-cell CDR3 sequences recovered (Supplemental Fig. S3). It should be noted that point-estimates of diversity or heterogeneity are only comparable within the subtype of interest, within a cohort. However, trends can be compared between point-estimates and patient cohorts. Furthermore, different cohorts based on their clinical context may show differences in immune cell infiltrates, even when considering simple cell marker differences between T-cells and B-cells. Interestingly, B-cell CDR3 recoveries dominated in the TCC cohort, accounting for an average of 93.41% (ranging from at least 53.24% to 99.95%). This contrasted with the CPTAC-3 renal cell carcinoma cohort, which contains generally less aggressive tumors, with only 40.9% of the cohort respectively comprised of stage 3 or 4 tumors.

Our results demonstrate that increased richness is indicative of larger and more advanced ccRCC tumors, which may be related to differences in underlying tumor biology. Evenness, as measured by the inflection point q, segregated patients based on survival. In a cross-validation cut-point analysis, patients with higher TRA evenness had a significantly improved overall survival compared to individuals with lower TRA evenness. These results indicate that patients’ TRA evenness, not richness, may be a possible prognostic biomarker and could have direct therapeutic consequences for response to systemic agents that elicit their effect in the tumor microenvironment (39). Furthermore, this evenness metric could be extended to other solid tumor types, as a quantitative metric of host contributions to immune infiltration that is a result of tumor evolution. These characterizations might become especially interesting in the context of terminally exhausted CD8+ T cells, which were recently shown to be enriched in advanced renal cell carcinoma, interacting with M2-like tumor-associated macrophages leading to immune dysfunction and poorer prognosis (44).

We identified demographic differences based on the CDR3 diversity of B-cell receptors, both individually and in combination, showing increased high-q diversity. This amounted to decreased dominance of the most abundant sequence, in non-white individuals compared to white individuals in the Moffitt TCC cohort. However, in this cohort, 88.5% of the patients were white (Table 1) and we were unable to confirm these results in the CPTAC-3 cohort due to missing data. Nevertheless, previously found race-related differences in BCR pathway activation in African Americans compared to European Americans lends support to our finding in differences in BCR dominance/clonality diversity (45). Furthermore, high-q diversity/sequence dominance may reveal gender-based differences in the ccRCC microenvironment. We found that females had higher clonality compared to male patients, which persists in the CPTAC-3 cohort. These demographic differences need to be further investigated, but our results suggest underlying race and gender differences in the heterogeneity of ccRCC microenvironments as reflected in tumor immune infiltration differences.

Biodiversity has historically been summarized into: alpha-diversity, which measures a single community’s diversity; beta-diversity, which quantifies the relative change of species between communities; and gamma diversity, which measures the total diversity in ecology (46). Diversity at the individual receptor subtype-level relates closest to alpha-diversity, which is then compared across the patients in a cohort. In a beta-diversity context, we see that many of the trends hold true between the Moffitt TCC and CPTAC-3 cohorts. The most commonly used diversity measures applied to cancer systems have been Shannon and Simpson indices (47, 48), which are special cases of the GDI at intermediate values of the parameter *q* (49, 50). Our analysis found that it is at the extremes of the continuum of diversity measures (low-q and high-q values) that we can stratify patients in clinically meaningful ways (for example, Fig. 2 vs. Supplemental Fig. S6). In this sense, novel properties of the GDI that are discussed here may allow a more nuanced, and thus more clinically comprehensive characterization of sequence heterogeneity. These novel objective diversity scales could have important applications for other systems in which tumor heterogeneity with its ecological and evolutionary impact is quantified.

Different point-estimates based on generalized diversity give unique information about the tumor. Increased richness in TRA and IGL diversity informs the size and aggressiveness of a tumor. Dominance of the most abundant sequence segregates patients based on prognosis. We identified a novel measure of evenness among immune receptor subtypes that could accurately classify patients’ overall survival. We also found important differences in receptor subtype contributions based on patient demographics such as race and gender. Using these diversity metrics, we identify a new statistical approach to stratify ccRCC patients-based differences in immune infiltration diversity and further guide precision oncology.

## Data Availability

Code for VDJ epitope recoveries from patient sequencing data (BAM files) is publicly available on Docker at https://hub.docker.com/r/bchobrut/vdj and GitHub at https://github.com/bchobrut/vdj_recovery.
Code for calculating patient CDR3 diversity from CDR3 recoveries is publicly available at: https://github.com/mcfefa/OncoDiversity.jl. The code and documentation describing how to run our pipeline and reproduce our results are open-source and publicly available through the OncoDiversity.jl GitHub repository. A virtual machine producing the full diversity environment is available on Code Ocean: https://codeocean.com/capsule/9959428/tree/v1

https://codeocean.com/capsule/9959428/tree/v1

## ABBREVIATIONS

CDR3: complementarity determining region-3
GDI: generalized diversity index
IP: inflection point
KM: Kaplan-Meier survival curve
OS: overall survival
RFS: recurrence free survival
TCR: T-cell receptor
TRA: T-cell receptor alpha
TRB: T-cell receptor beta
TRG: T-cell receptor gamma
TRD: T-cell receptor delta
BCR: B-cell receptor
IGH: B-cell immunoglobulin heavy chain
IGK: B-cell immunoglobulin kappa light chain
IGL: B-cell immunoglobulin lambda light chain

## DATA AND CODE AVAILABILITY

Code for VDJ epitope recoveries from patient sequencing data (BAM files) is publicly available on Docker at https://hub.docker.com/r/bchobrut/vdj and GitHub at https://github.com/bchobrut/vdj_recovery.

Code for calculating patient CDR3 diversity from CDR3 recoveries is publicly available at: https://github.com/mcfefa/OncoDiversity.jl. The code and documentation describing how to run our pipeline and reproduce our results are open-source and publicly available through the OncoDiversity.jl GitHub repository. A virtual machine producing the full diversity environment is available on Code Ocean (DOI: 10.24433/CO.1660327.v1).

## AUTHOR CONTRIBUTIONS

MCF: conceptualization, data curation, formal analysis, investigation, methodology, software, validation, visualization, writing.

NC: data curation, critical review.

BIC: data curation, adaptive immune receptor recombination read extractions from the Moffitt TCC cohort RNAseq files.

YK: statistical analysis, validation, critical review.

JKT: logistic support, data curation, statistical analysis, critical review.

AB: logistic support, data curation, statistical analysis, critical review.

JJM: investigation, critical review.

MF: logistic support, data curation.

ES: logistic support, data curation.

JD: critical review.

SSAF: data curation, adaptive immune receptor recombination read extractions from the Moffitt TCC cohort RNAseq files.

JFA: data curation, adaptive immune receptor recombination read extractions from the Moffitt TCC cohort RNAseq files.

ENK: logistic support, data curation.

GB: adaptive immune receptor recombination read extractions from the Moffitt TCC cohort RNAseq files, critical review, study supervision.

BJM: conceptualization, investigation, methodology, validation, critical review, writing, study supervision.

PMA: conceptualization, investigation, methodology, software, validation, critical review, writing, study supervision.

## ACKNOWLEDGEMENTS

This work was supported in part by the Biostatistics and Bioinformatics Shared Resource at the H. Lee Moffitt Cancer Center & Research Institute, a National Cancer Institute–designated Comprehensive Cancer Center (P30-CA076292). Total Cancer Care (TCC) Protocol at Moffitt Cancer Center, which was enabled in part by the generous support of the DeBartolo Family. Some data used in this publication were generated by the National Cancer Institute Clinical Proteomic Tumor Analysis Consortium (CPTAC), available to which author via dbGaP project approval number 6757. Editorial assistance was provided by the Moffitt Cancer Center’s Scientific Editing Department by Dr. Paul Fletcher and Daley Drucker (no compensation was given beyond their regular salaries). We also thank Gregory J. Kimmel, PhD for valuable comments.

---

**Supplementary Figure S1:**
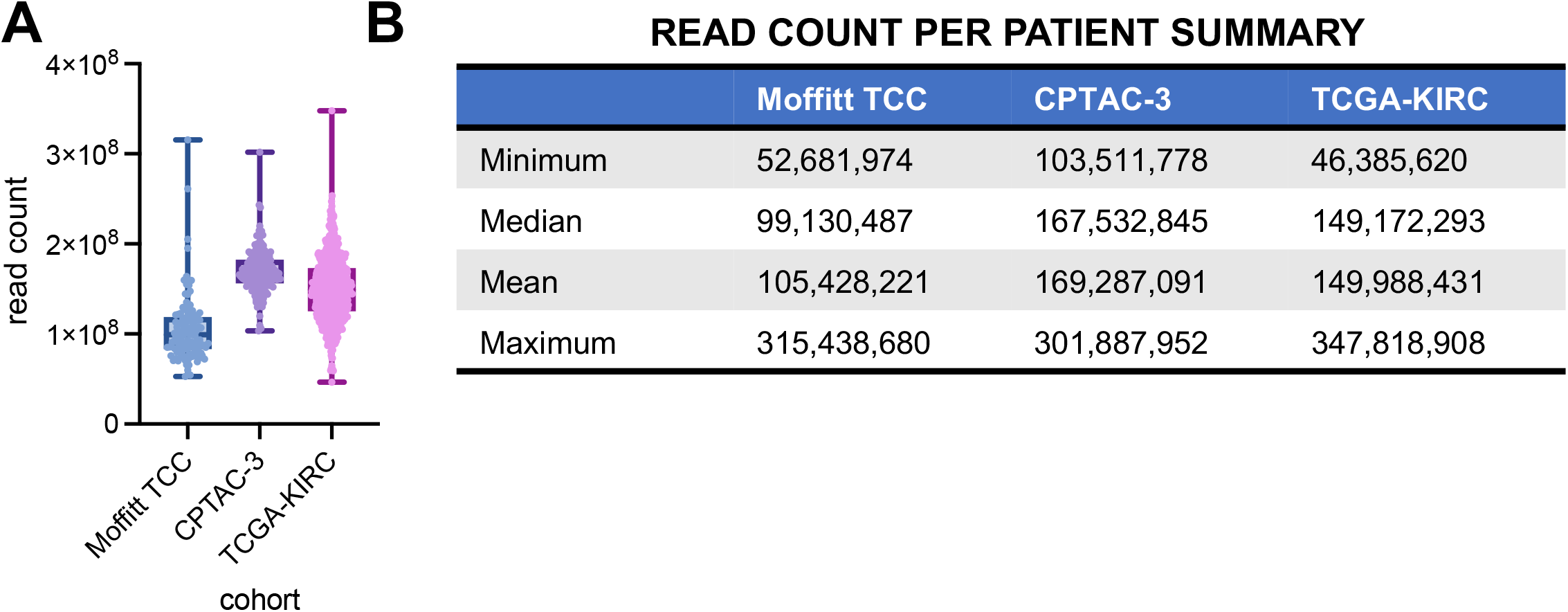
Distributions of read counts per sample across all three cohorts. (A) Distribution of individual reads counts per patient across the Moffitt TCC Cohort, CPTAC-3 Cohort, and the TCGA-KIRC Cohort. (B) Summary table of the minimum, median, mean, and maximum number of read counts across all three cohorts.

**Supplementary Figure S2:**
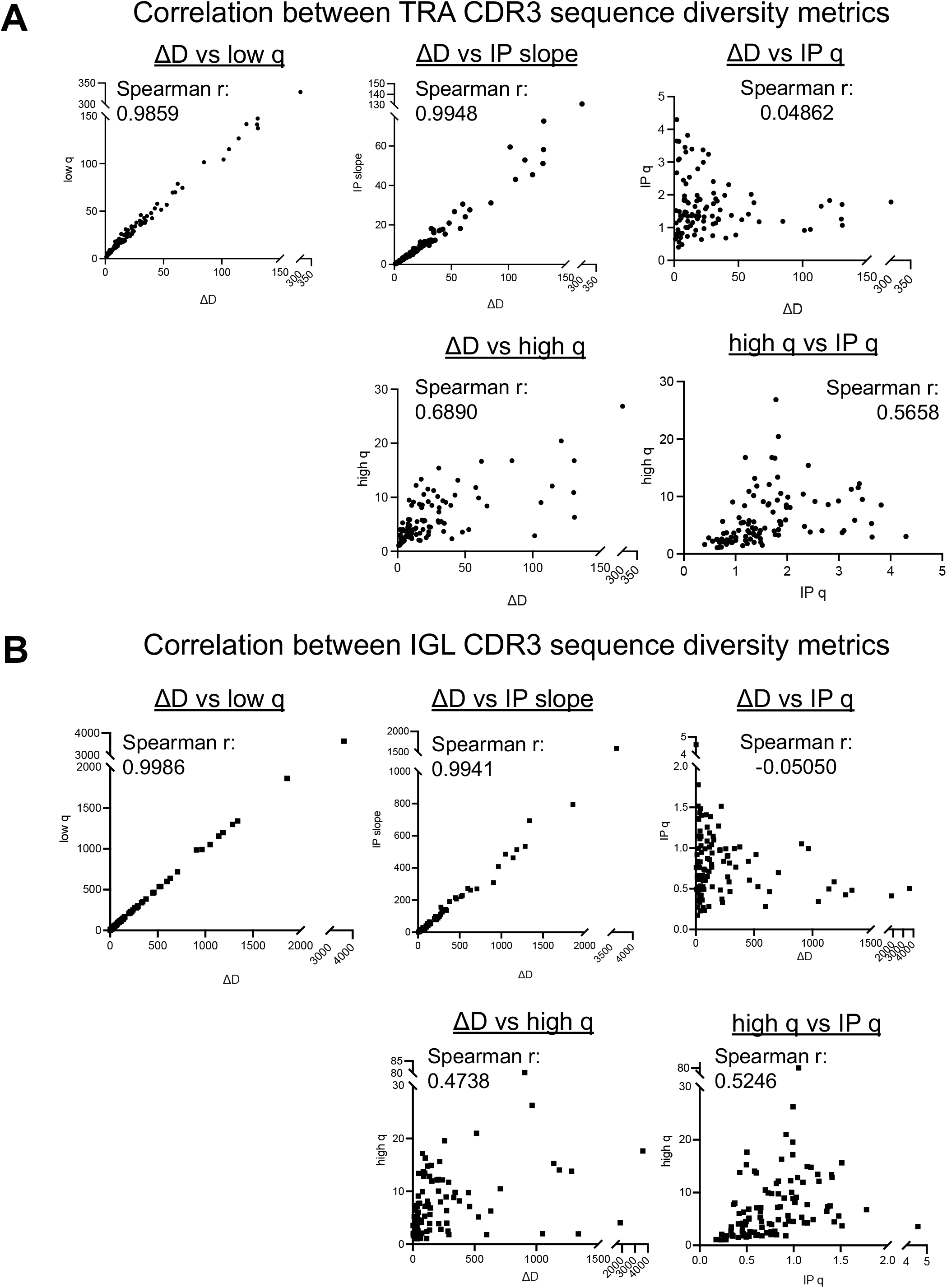
Correlation analysis showing the low q, ΔD, and inflection point slope diversity metrics are highly correlated and give the same information. Correlations were assessed using a Spearman correlation coefficient (r) across all samples in the cohort segregated by receptor type, highlighting TRA & IGL CDR3 sequence diversities here.

**Supplemental Figure S3:**
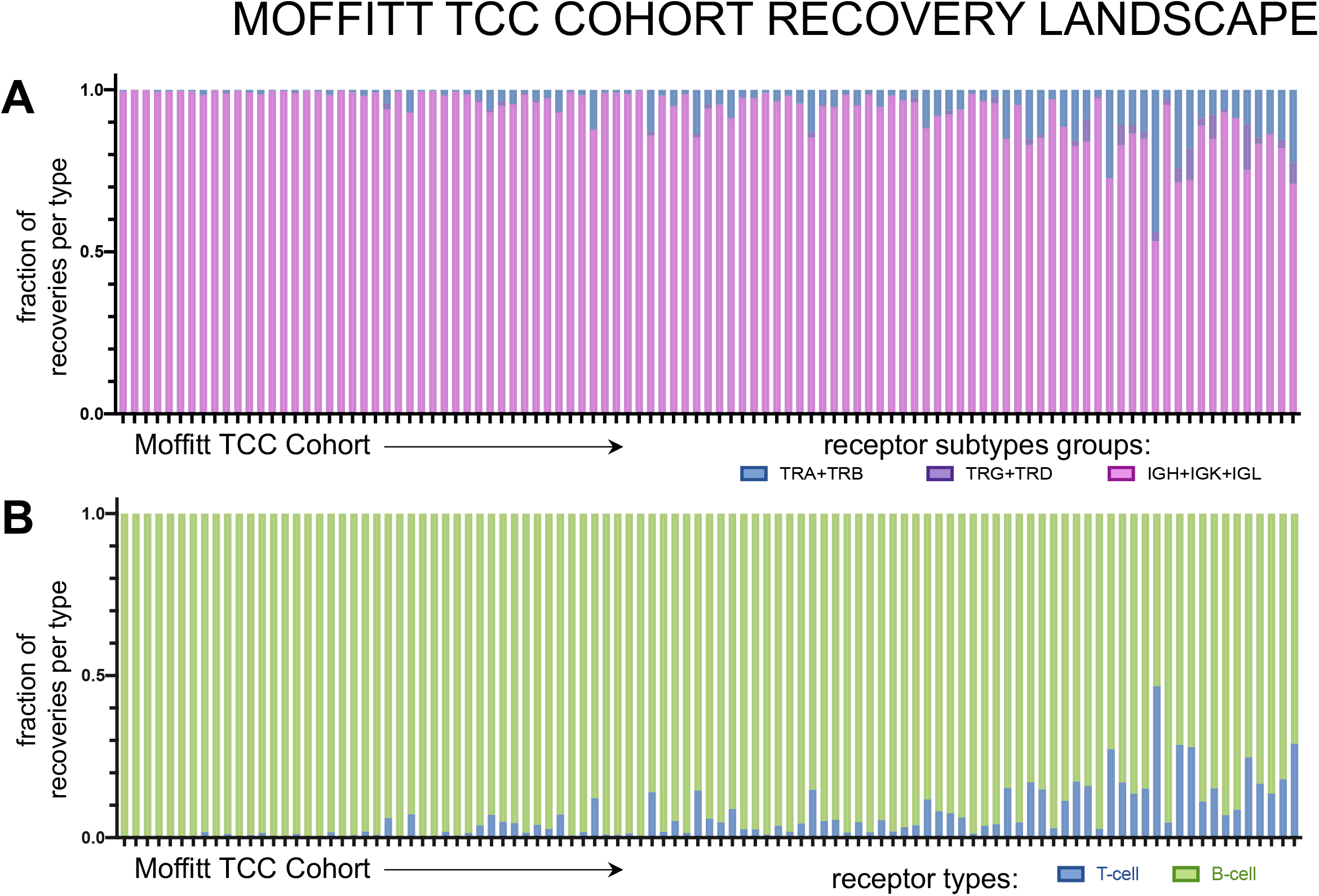
Tumor infiltrating lymphocyte receptor recovery landscape the Moffitt TCC Cohort of clear cell renal cell carcinoma. Patient tumors undergo bulk RNA sequencing and then CDR3 sequences from TCR and BCR receptors were recovered. Then for each patient, CDR3s are segregated by receptor class. Fig. 1B shows total recoveries per patient was reported in a bar plot with each bar reflecting the total number of recoveries from each patient and the proportion of each recovery type per patient was reported for individual T-cell and B-cell receptor types. The proportion of each recovery type per patient grouped by common receptor combinations (C) and per cell type combinations (D) are reflected in the bar charts above.

**Supplemental Figure S4:**
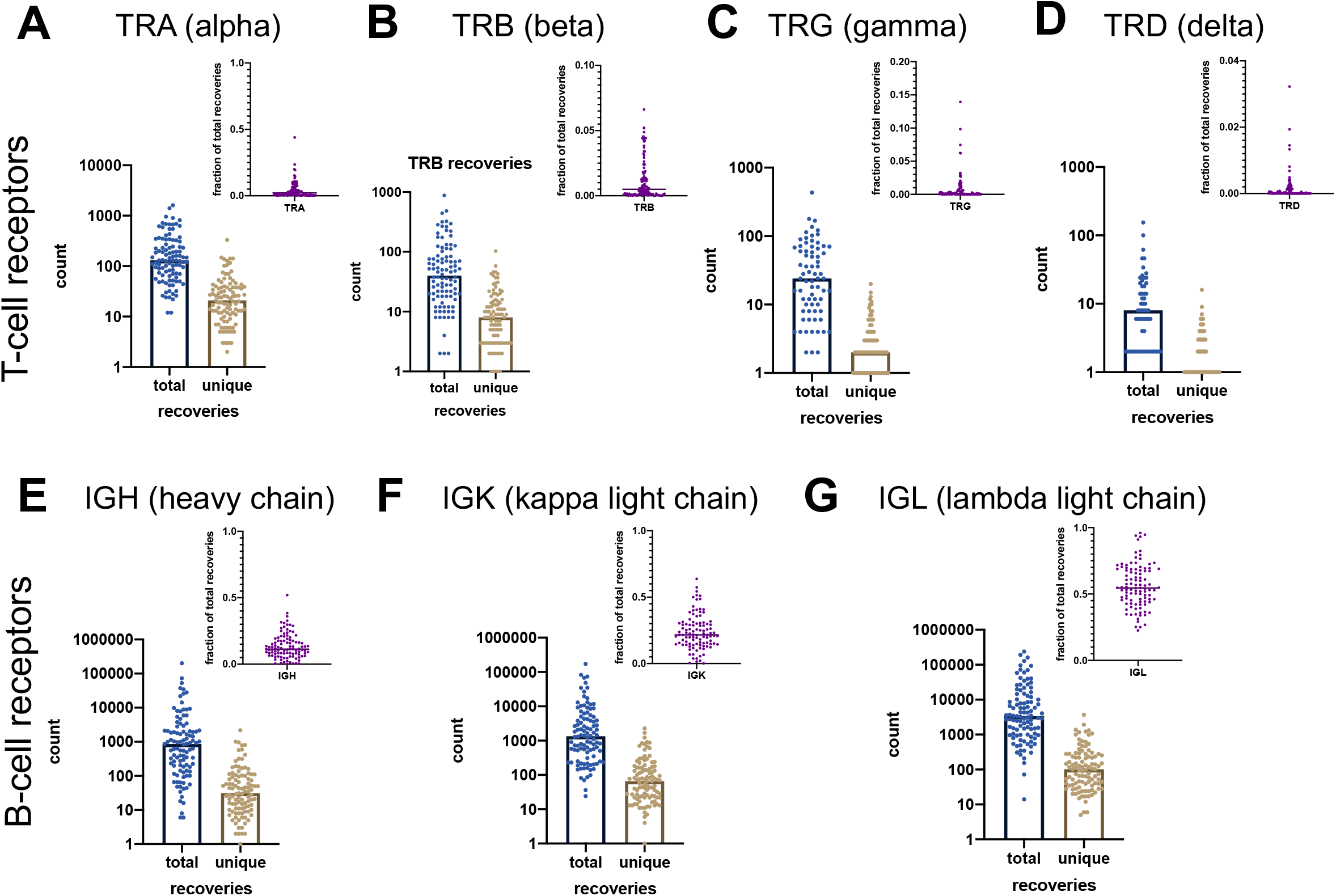
Recovery counts per patient across all seven tumor-infiltrating lymphocyte receptor subtypes in the Moffitt TCC Cohort of clear cell renal cell carcinoma patients. Patient tumors undergo bulk RNA sequencing and then CDR3 sequences from TCR and BCR receptors were recovered. Then for each patient, CDR3 sequences were segregated by receptor class. Total and unique recovery counts per patient as well as the fraction of those recoveries out of all recoveries for each patient were reported for T-cell receptors (TR) type (A) alpha, (B) beta, (C) gamma, and (D), delta, as well as B-cell receptors immunoglobulin (IG) type (E) heavy chain, (F) kappa light chain, and (G) lambda light chain.

**Supplementary Figure S5.**
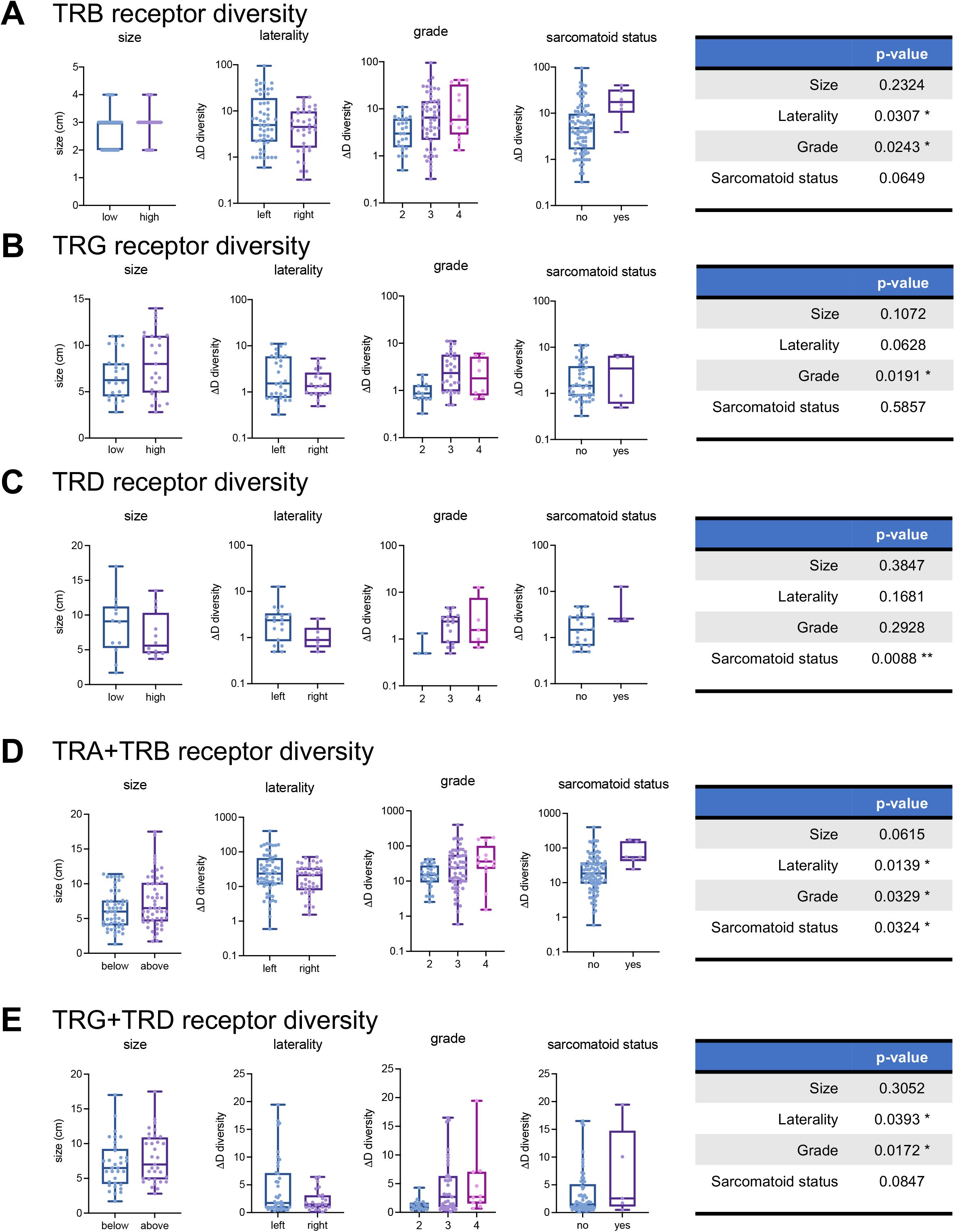
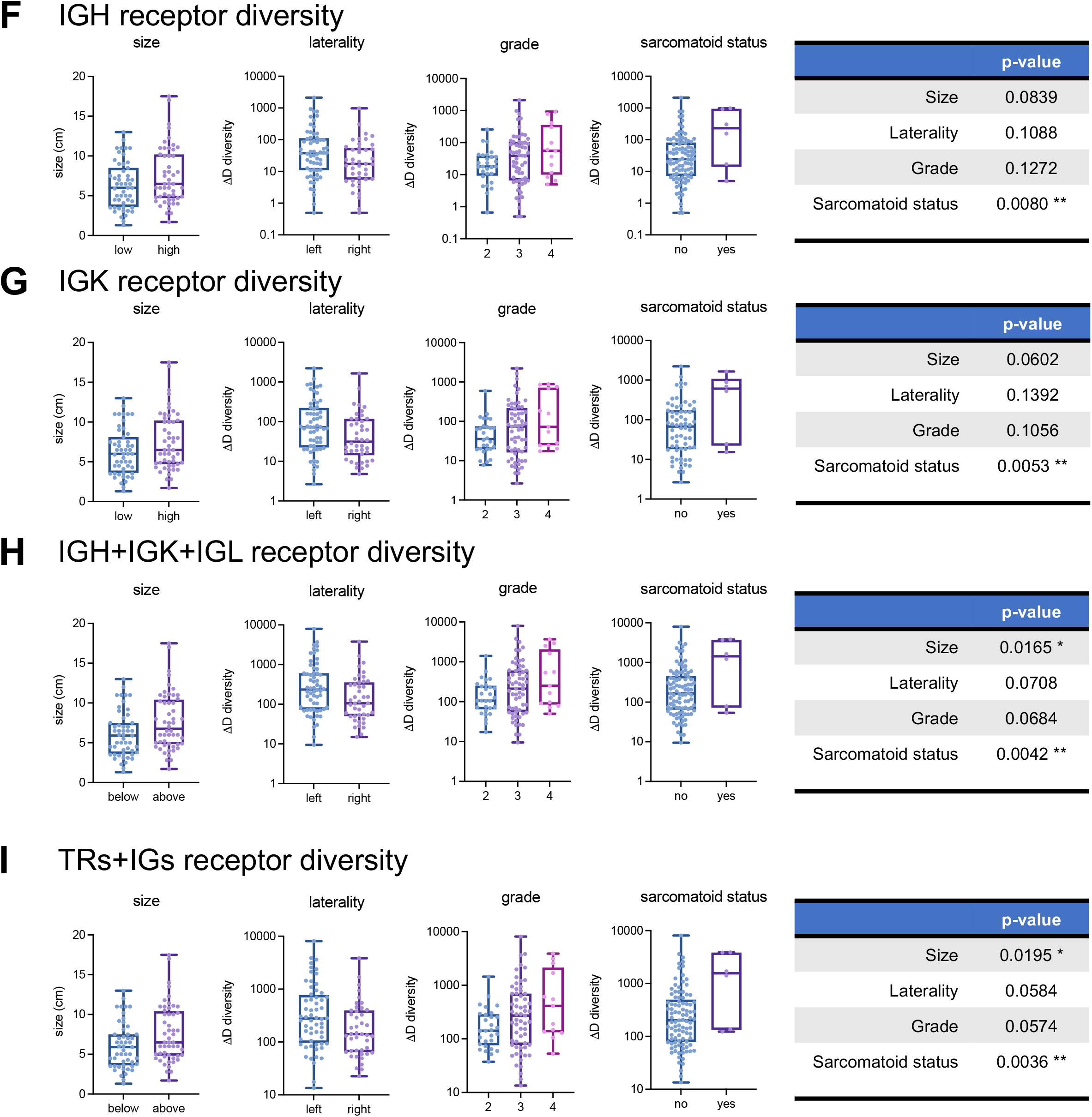
Species richness associated with larger and worse tumors across other receptor combinations. Receptor diversity with respect to largest diameter (size, cm), laterality, grade, and sarcomatoid status for (A) TRB recoveries, (B) TRG recoveries, (C) TRD recoveries, (D) TRA and TRB recoveries, (E) TRG and TRD recoveries, (F) IGH recoveries, (G) IGK recoveries, (H) IGH, IGK, IGL (or B-cell) recoveries, and (I) total (TRs+IGs) recoveries. Similar plots for TRA recoveries and IGL recoveries are in Fig 2. p-value significance represented by * < 0.05, ** < 0.01, *** < 0.001

**Supplemental Figure S6:**
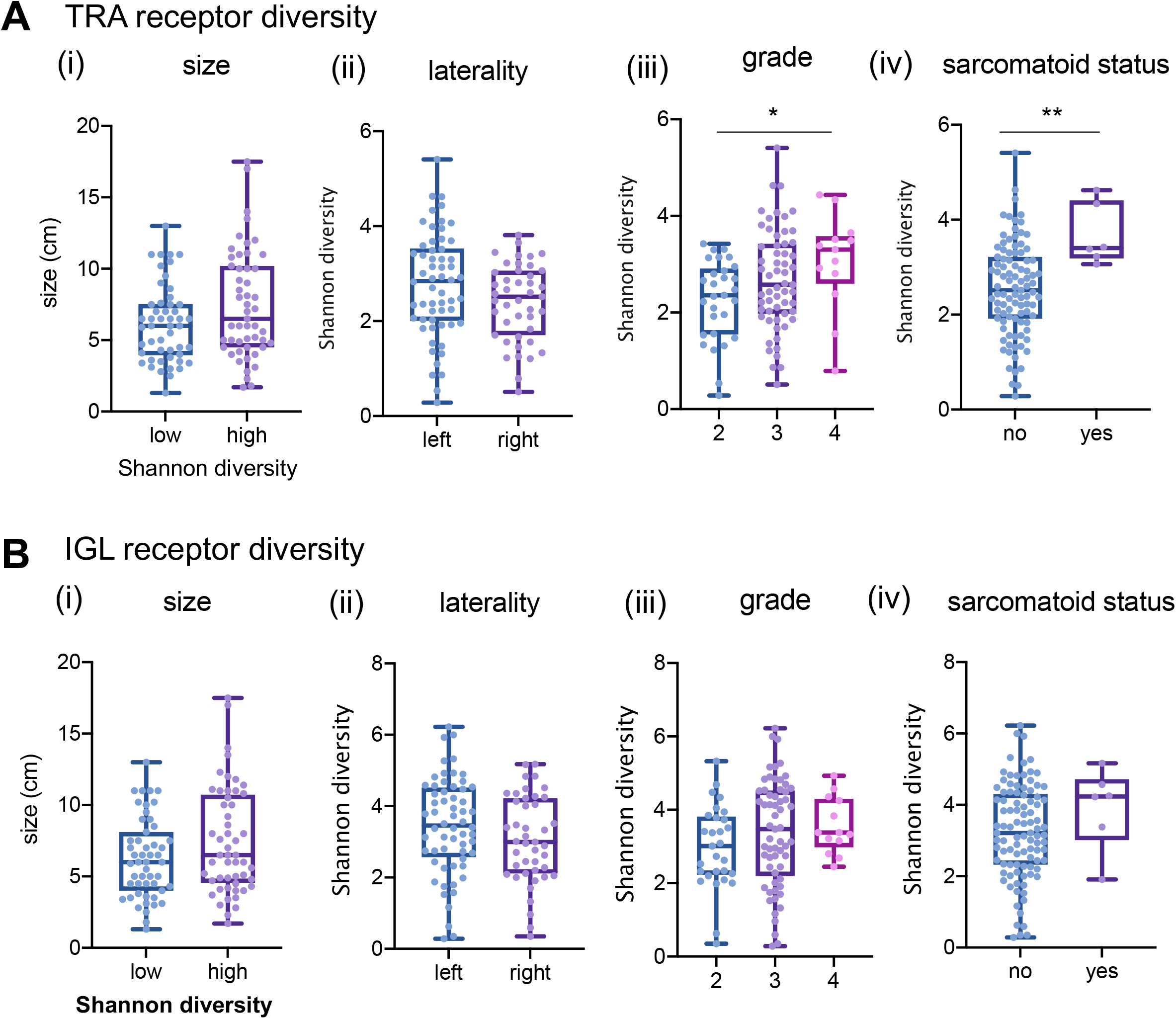
Shannon diversity does not capture the same clinical information in TRA and IGL receptors. (A) TRA receptor CDR3 sequence Shannon diversity across the Moffitt TCC Cohort trends in (i) largest diameter tumors (low diversity had mean diameter of 6.2 cm and high diversity had a mean diameter for 7.5 cm; p-value: 0.0506), (ii) tumor laterality (score of 2.762 on left vs 2.401 on right; p-value: 0.0729), (iii) grade (mean score from grade 2 was 2.262, mean score from grade 3 was 2.682, and mean score from grade 4 was 3.049; p-value: 0.0414), and (v) sarcomatoid status (no mean score of 2.541 vs yes mean score of 3.675; p-value 0.0065). (B) IGL receptor CDR3 sequence Shannon diversity no significant trends for (i) size (low diversity had mean diameter of 6.3 cm and high diversity had a mean diameter for 7.5 cm; p-value: 0.0676), (ii) laterality (score of 3.436 vs 3.063; p-value: 0.1548), (iii) grade (mean score from grade 2 was 2.979, mean score from grade 3 was 3.363, and mean score from grade 4 was 3.564; p-value: 0.3060), and (iv) sarcomatoid status (no had a mean score of 3.241 vs yes with a mean score of 3.917; p-value: 0.2216). Unpaired t-tests were used to compare two group data and ANOVA was used to compare grade, three group data. p-value significance represented by * < 0.05, ** < 0.01, *** < 0.001

**Supplemental Figure S7:**
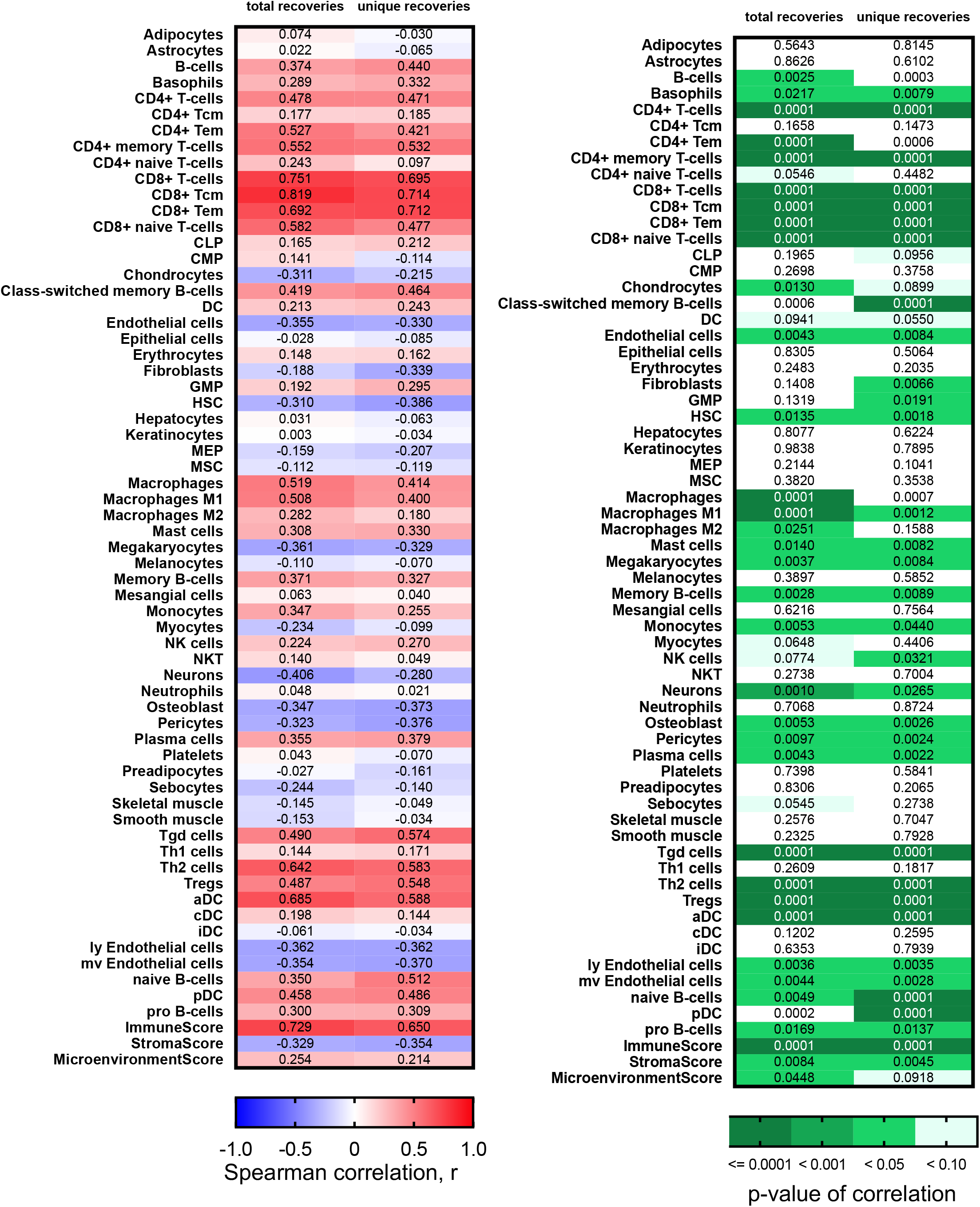
TRA receptor full xCELL score Spearman correlation heatmaps and associated p-values.

**Supplemental Figure S8:**
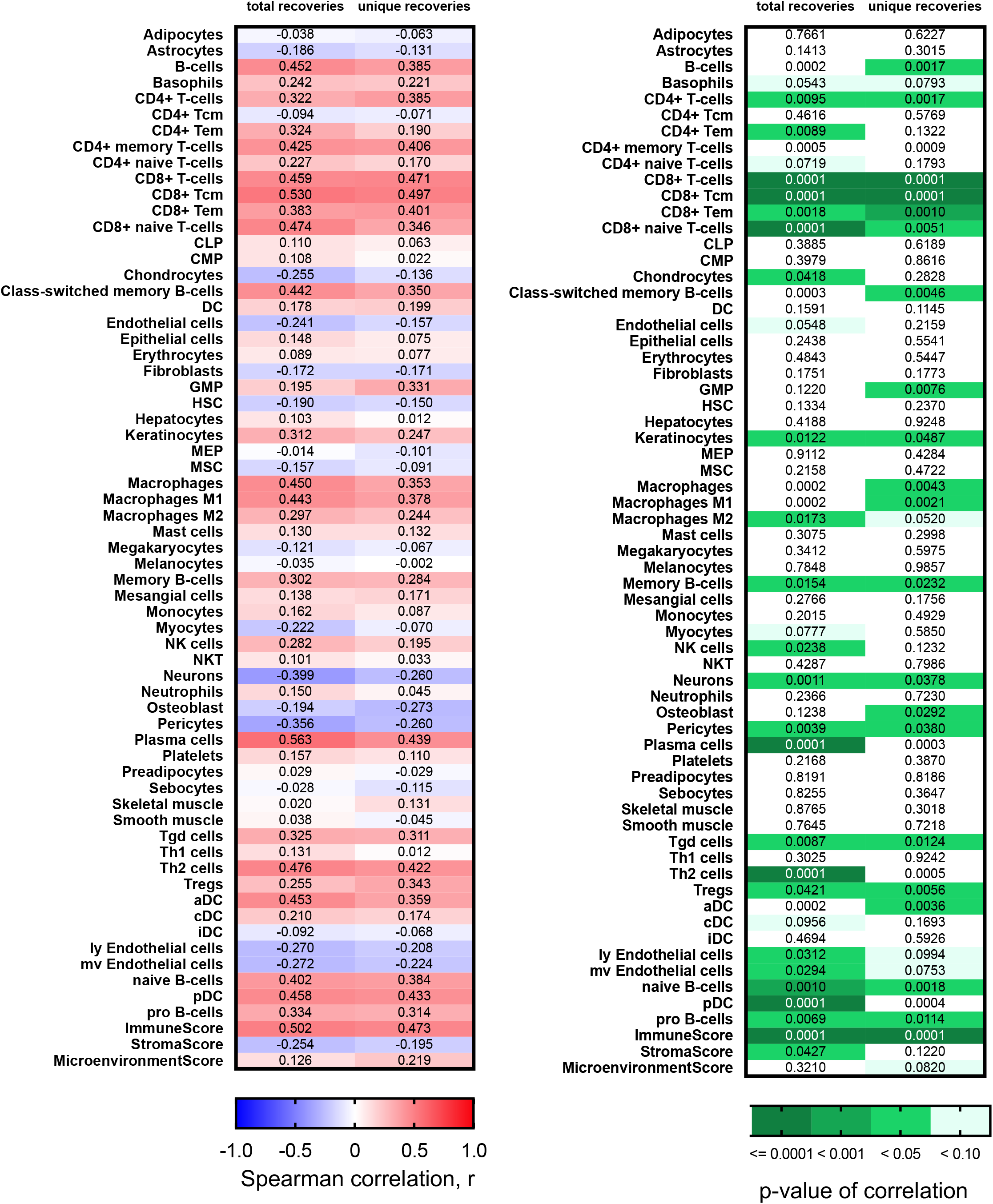
IGL receptor full xCELL score Spearman correlation heatmaps and associated p-values.

**Supplemental Figure S9:**
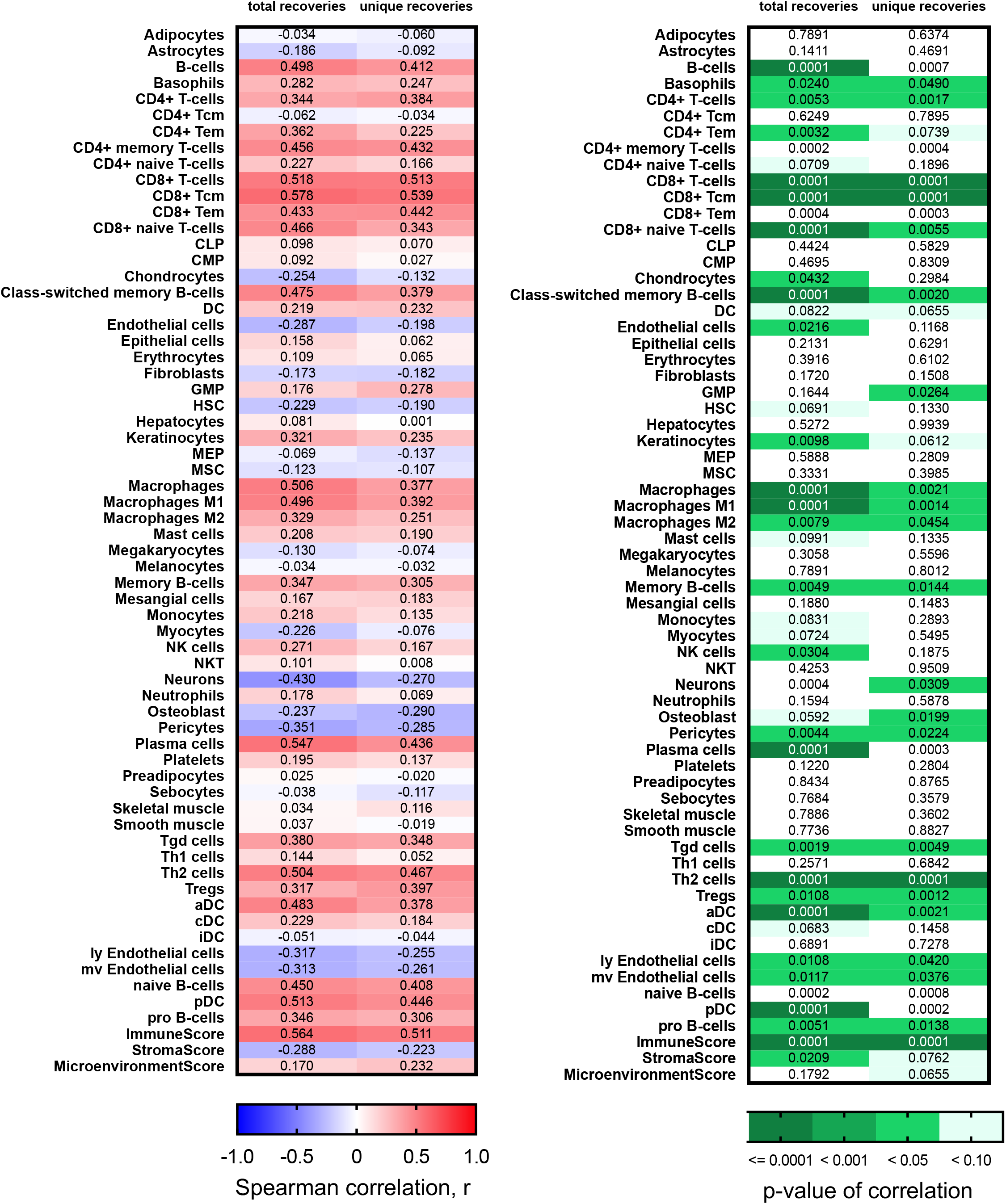
TRs+IGs receptors full xCELL score Spearman correlation heatmaps and associated p-values.

**Supplemental Figure S10:**
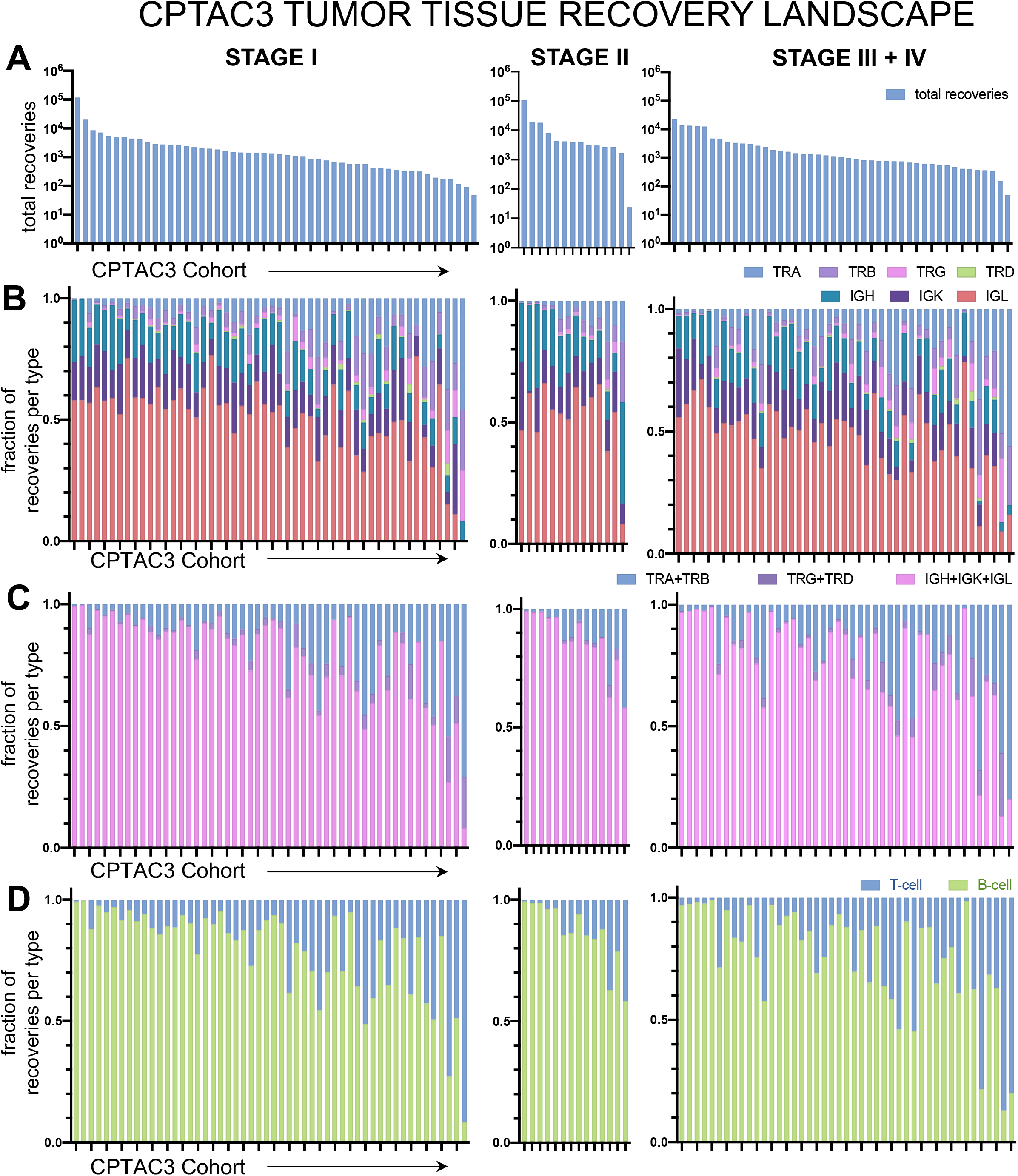
Tumor infiltrating lymphocyte receptor recovery landscape the tumor tissue from the CTPAC3 clear cell renal cell carcinoma cohort. Patient tumors undergo bulk RNA sequencing and then CDR3 sequences from TCR and BCR receptors were recovered. Then for each patient, CDR3s are segregated by receptor class. (A) Total recoveries per patient was reported in a bar plot with each bar reflecting the total number of recoveries from each patient. The proportion of each recovery type per patient was reported for individual T-cell and B-cell receptor types (B), common receptor combinations (C), and per cell type combinations (D).

**Supplemental Figure S11:**
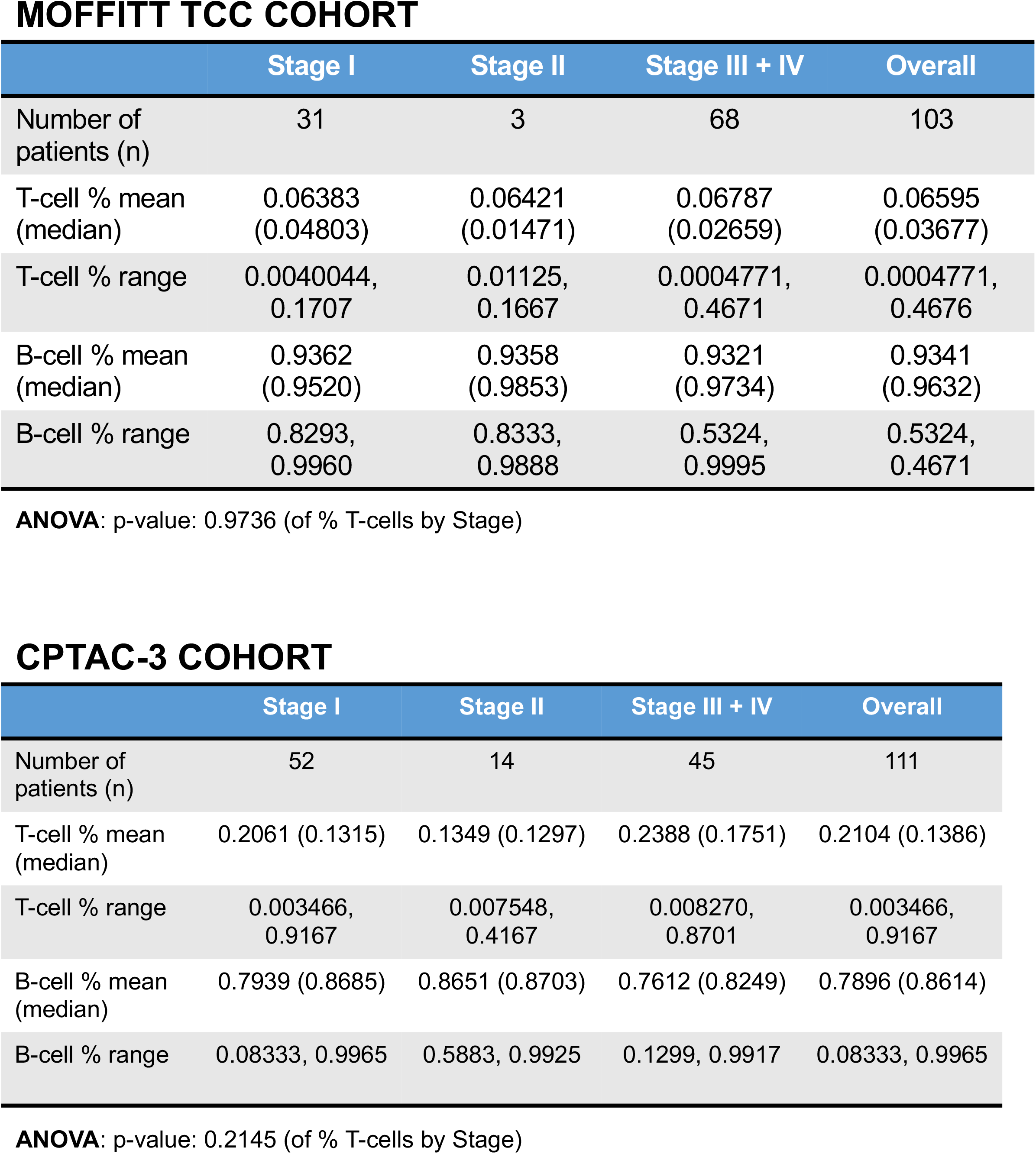
Comparison of T-cell and B-cell contributions to recovery landscape in the Moffitt TCC and CTPAC-3 clear cell renal cell carcinoma cohorts.

**Supplemental Figure S12:**
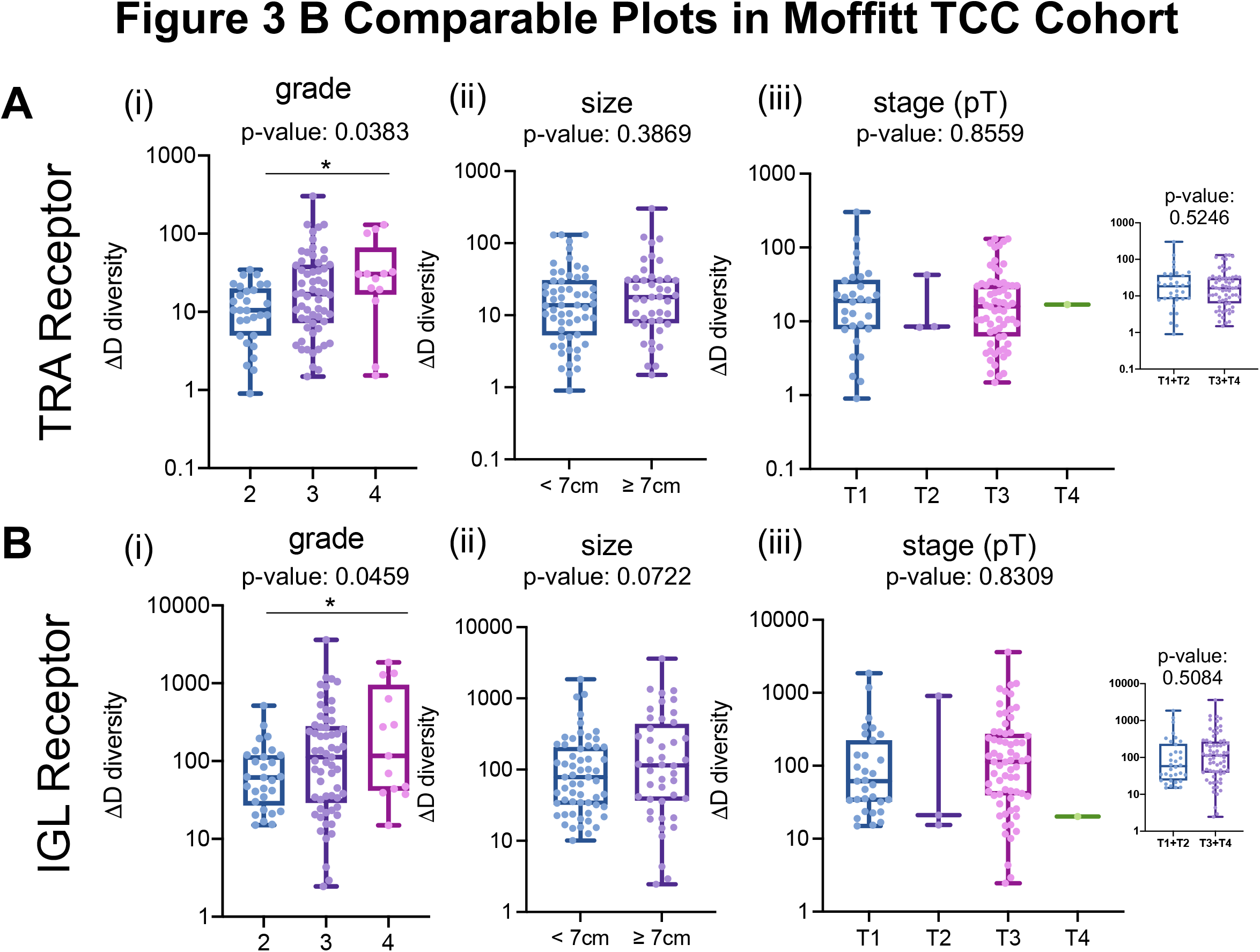
Comparable size, grade, and stage (pT) plots of ΔD diversity in TRA and IGL receptors of the Moffitt TCC cohort. Based on the clinical data available, some of the comparisons between the Moffitt TCC cohort and CPTAC-3 cohort are a little different. The panels in this figure show the same grouping of data from the Moffitt TCC cohort as shown in the validation CPTAC-3 cohort data shown in Fig. 3B. (A) TRA receptor CDR3 sequence ΔD diversity was increased diversity in (i) with higher grade tumors (replicate of figure panel Fig. 2A iii), (ii) larger diameter tumors (tumors with a largest tumor diameter above 7cm had a mean ΔD diversity of 32.67, tumors with a diameter below 7cm had a mean ΔD diversity of 25.47; p-value: 0.03869), and (iii) stage (mean score for T1 tumors was 33.84, mean score for T2 tumors was 19.77, mean score for T3 tumors was 27.11, and mean score for T4 tumors was 16.87, p-value 0.8559; mean score for T1+T2 tumors was 32.56 and for T3+T4 tumors was 26.95, p-value: 0.5246). (B) IGL receptor CDR3 sequence ΔD diversity showed similar trends as TRA receptor CDR3 sequence diversity for (i) grade (replicate of figure panel Fig. 2B iii), (ii) size (tumors with a largest tumor diameter above 7cm had a mean ΔD diversity of 349.0, tumors with a diameter below 7cm had a mean ΔD diversity of 179.4; p-value: 0.0722), and (iii) stage (mean score for T1 tumors was 198.8, mean score for T2 tumors was 314.1, mean score for T3 tumors was 279.3, and mean score for T4 tumors was 20.20, p-value 0.8309; mean score for T1+T2 tumors was 209.0 and for T3+T4 tumors was 275.5, p-value: 0.5084). Unpaired t-tests were used to compare two group data and ANOVA was used to compare grade, three group data. p-value significance represented by * < 0.05, ** < 0.01, *** < 0.001

**Supplemental Figure S13:**
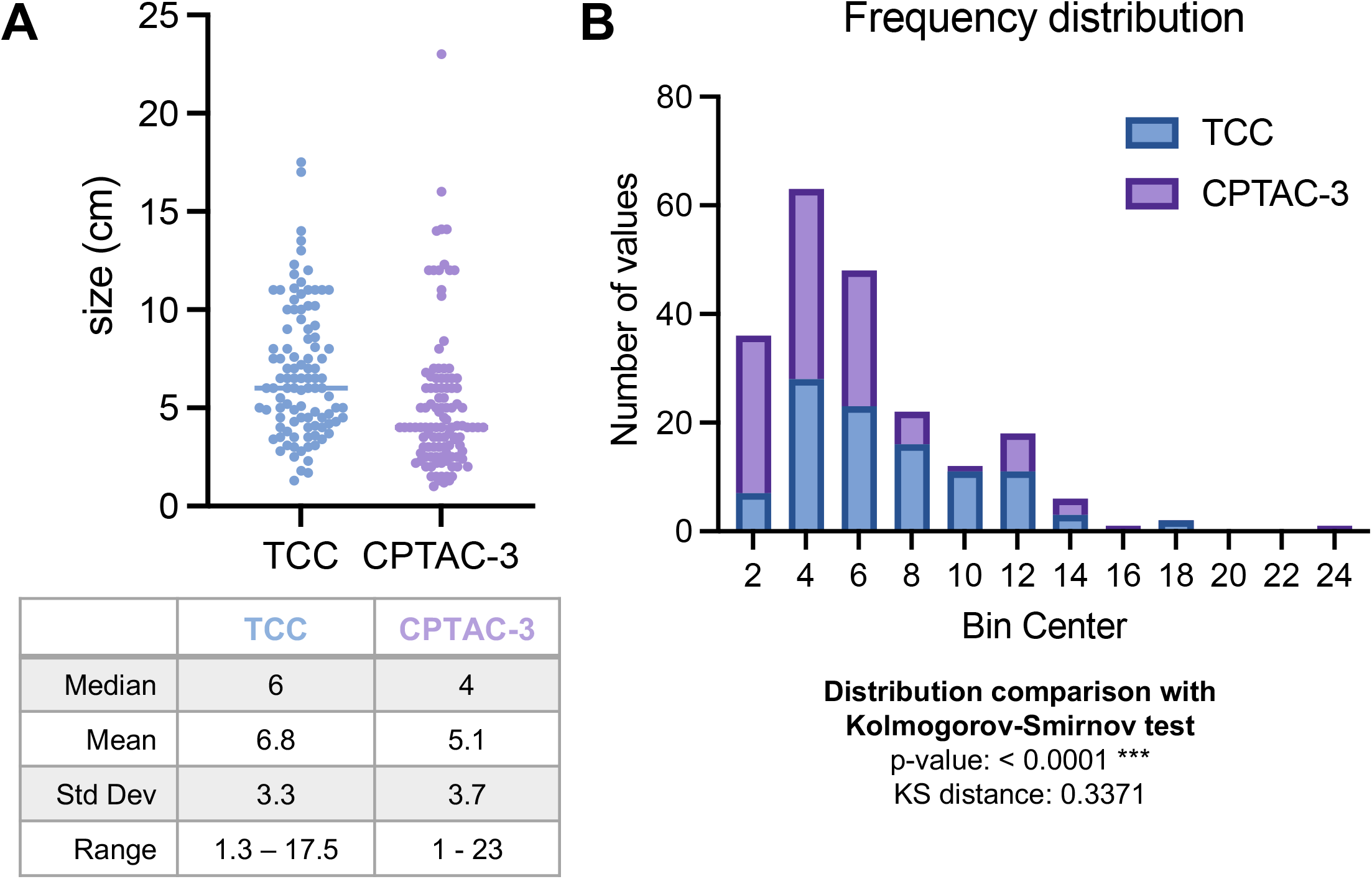
Comparison of the largest diameter size distribution of the Moffitt TCC and CPTAC-3 renal cell carcinoma cohorts. (A) Distribution of largest diameter sizes (in cm) of tumors from the Moffitt TCC and CPTAC-3 Cohort with descriptive statistics showing that mean and median diameter of CPTAC-3 tumors is smaller than the mean and median diameter of Moffitt TCC tumors. (B) Histogram of the frequency distribution of tumors that show the distribution of Moffitt TCC and CPTAC-3 tumors are statistically significantly different (Kolmogorov-Smirnov test, distance of 0.3371 and p-value < 0.0001).

**Supplemental Figure S14:**
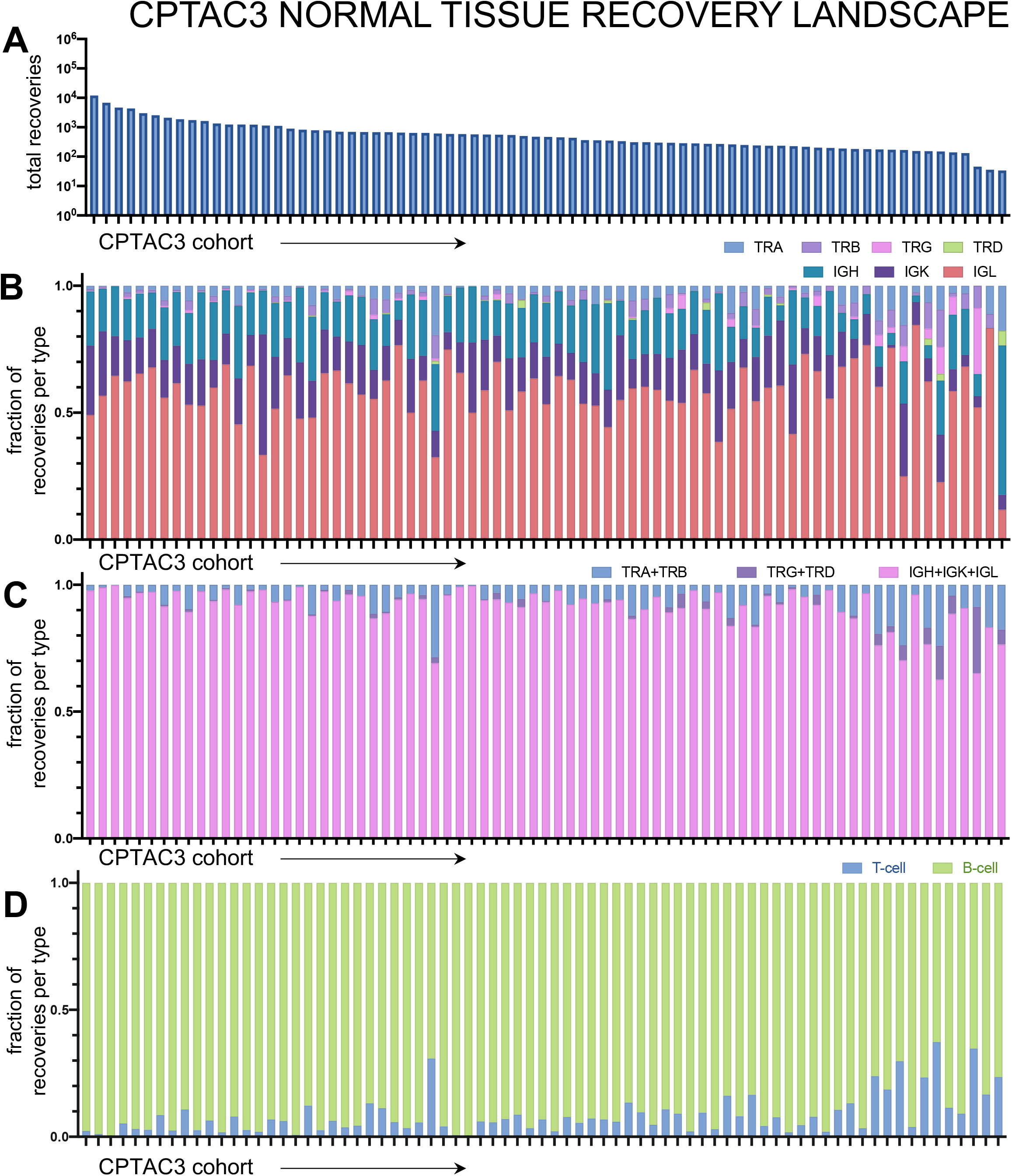
Tumor infiltrating lymphocyte receptor recovery landscape the normal tissue from CPTAC3 clear cell renal cell carcinoma cohort. Patient tumors undergo bulk RNA sequencing and then CDR3 sequences from TCR and BCR receptors were recovered. Then for each patient, CDR3s are segregated by receptor class. (A) Total recoveries per patient was reported in a bar plot with each bar reflecting the total number of recoveries from each patient. The proportion of each recovery type per patient was reported for individual T-cell and B-cell receptor types (B), common receptor combinations (C), and per cell type combinations (D).

**Supplemental Figure S15:**
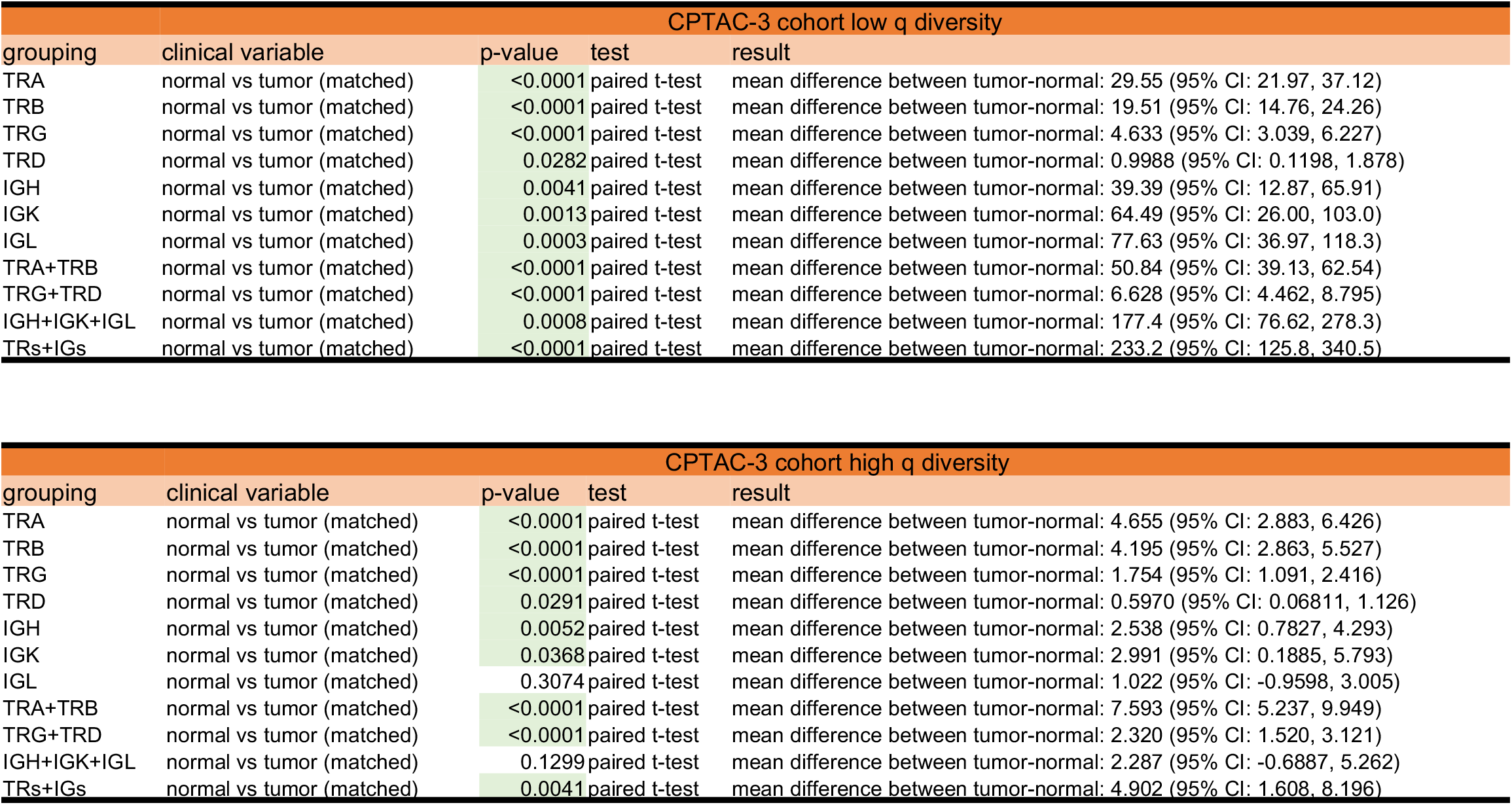
Statistical comparison of low q (richness) and high q (evenness) across all receptor subtypes and combinations in the CPTAC-3 cohort.

**Supplemental Figure S16:**
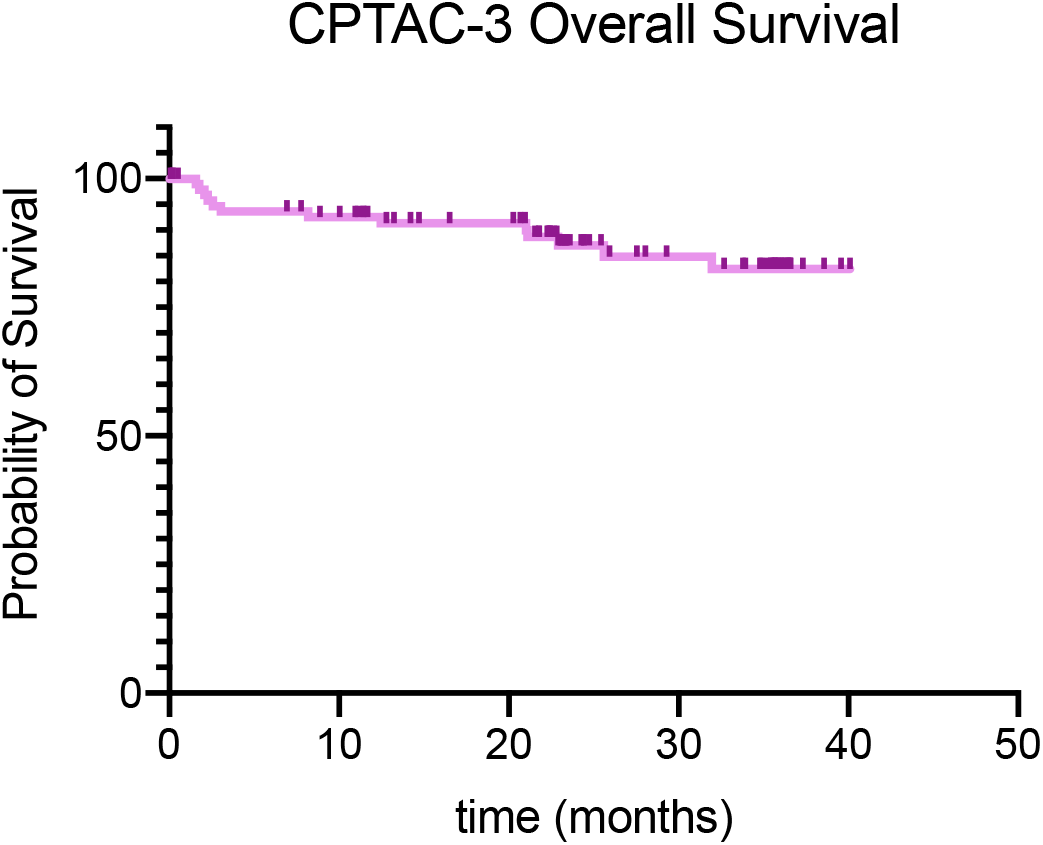
CPTAC-3 Cohort overall survival could not confirm the Moffitt TCC survival trend because over 85% of patients were censored.

**Supplemental Figure S17:**
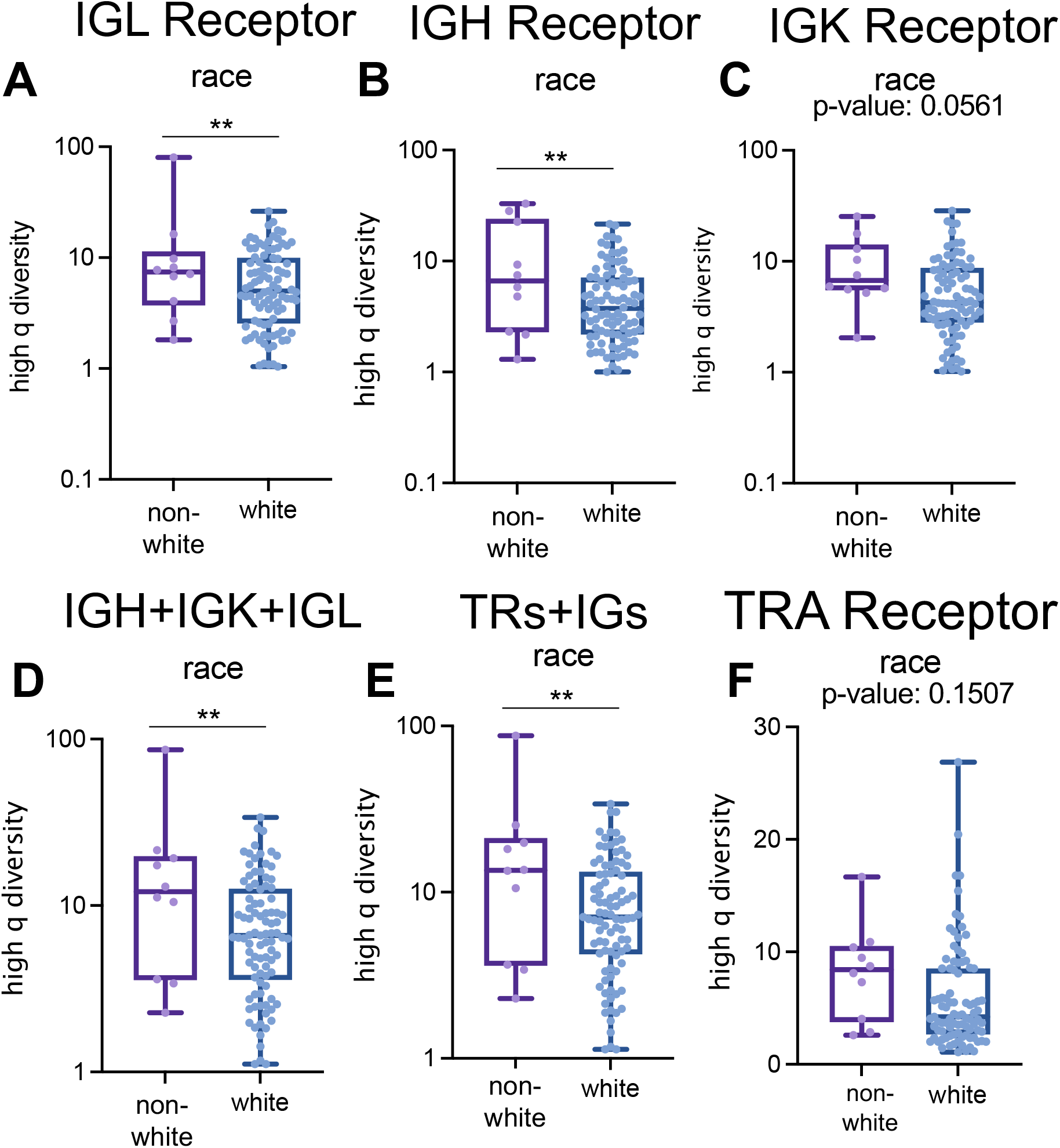
Generalized diversity index can capture race-based difference in B-cell receptor CDR3 recoveries in the Moffitt TCC cohort. High q diversity differences in the Moffitt TCC Cohort based on race in (A) IGL receptor (score of 6.883 in white vs score of 14.47 in non-white, p-value: 0.0095), (B) IGH receptor (score of 5.376 in white vs score of 11.69 in non-white, p-value: 0.0026), (C) IGK receptor (score of 6.282 in white vs score of 9.832 in non-white, p-value: 0.0561), (D) IGH+IGK+IGL receptors (score of 8.712 in white vs score of in non-white, p-value: 0.0026), (E) all receptors (score of 9.336 in white vs score of 19.67 in non-white, p-value: 0.0025), and (F) TRA receptor. Unpaired t-tests were used to compare categorical data. p-value significance represented by * < 0.05, ** < 0.01, *** < 0.001

**Supplemental Figure S18:**
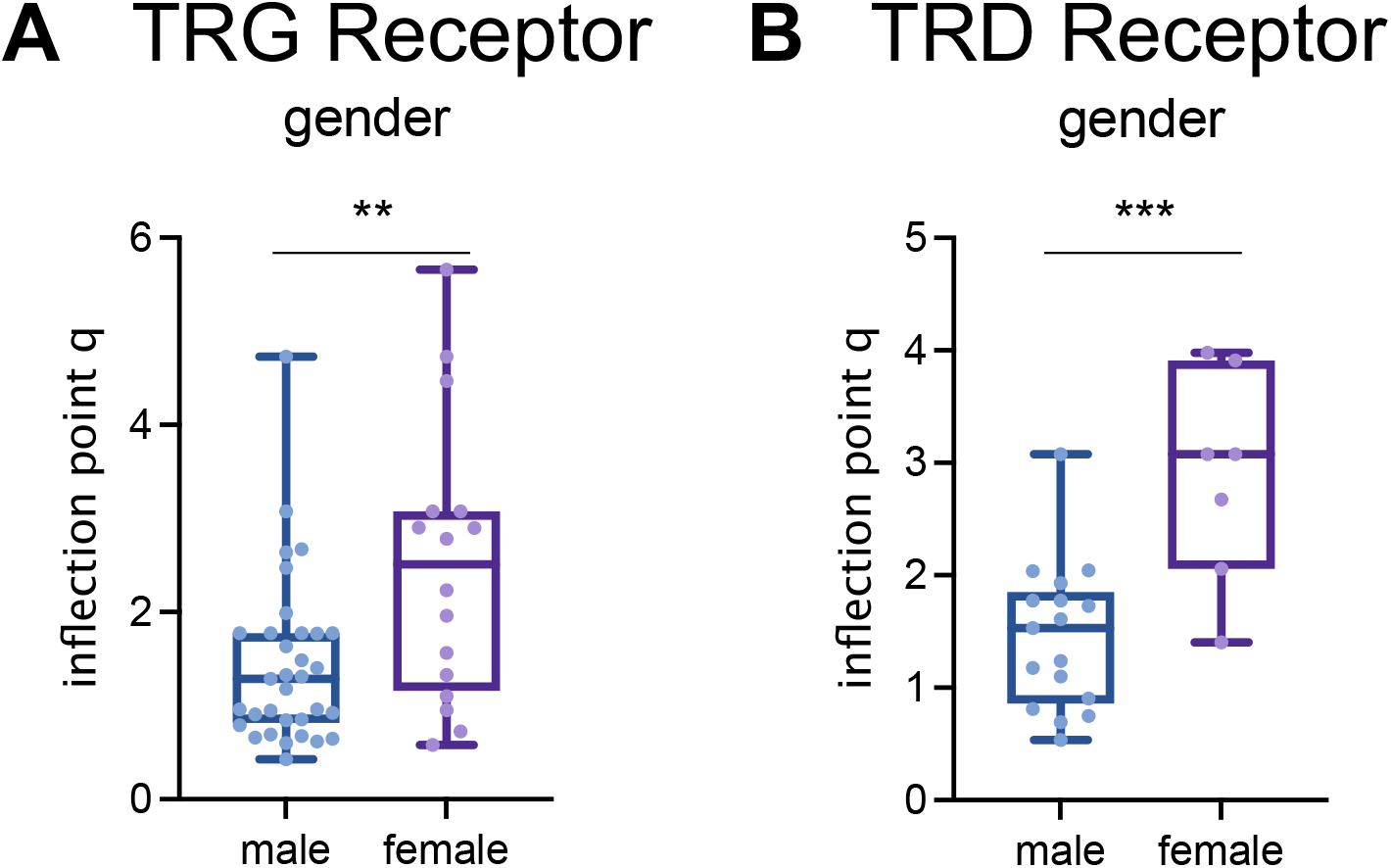
Inflection point q diversity can also capture gender-based difference in T-cell receptor CDR3 recoveries in the Moffitt TCC cohort. (A) T-cell receptor gamma (TRG) inflection point q (IP q) was elevated in female patients compared to males (mean IP q in male patients was 1.442, mean IP q in female patients was 2.503; p-value: 0.0033). (B) T-cell receptor delta (TRD) inflection point q was elevated in female patients compared to males (mean IP q in male patients was 1.454, mean IP q in female patients was 2.883; p-value: 0.0003). Unpaired t-tests were used to compare categorical data. p-value significance represented by * < 0.05, ** < 0.01, *** < 0.001

**Supplemental Figure S19:**
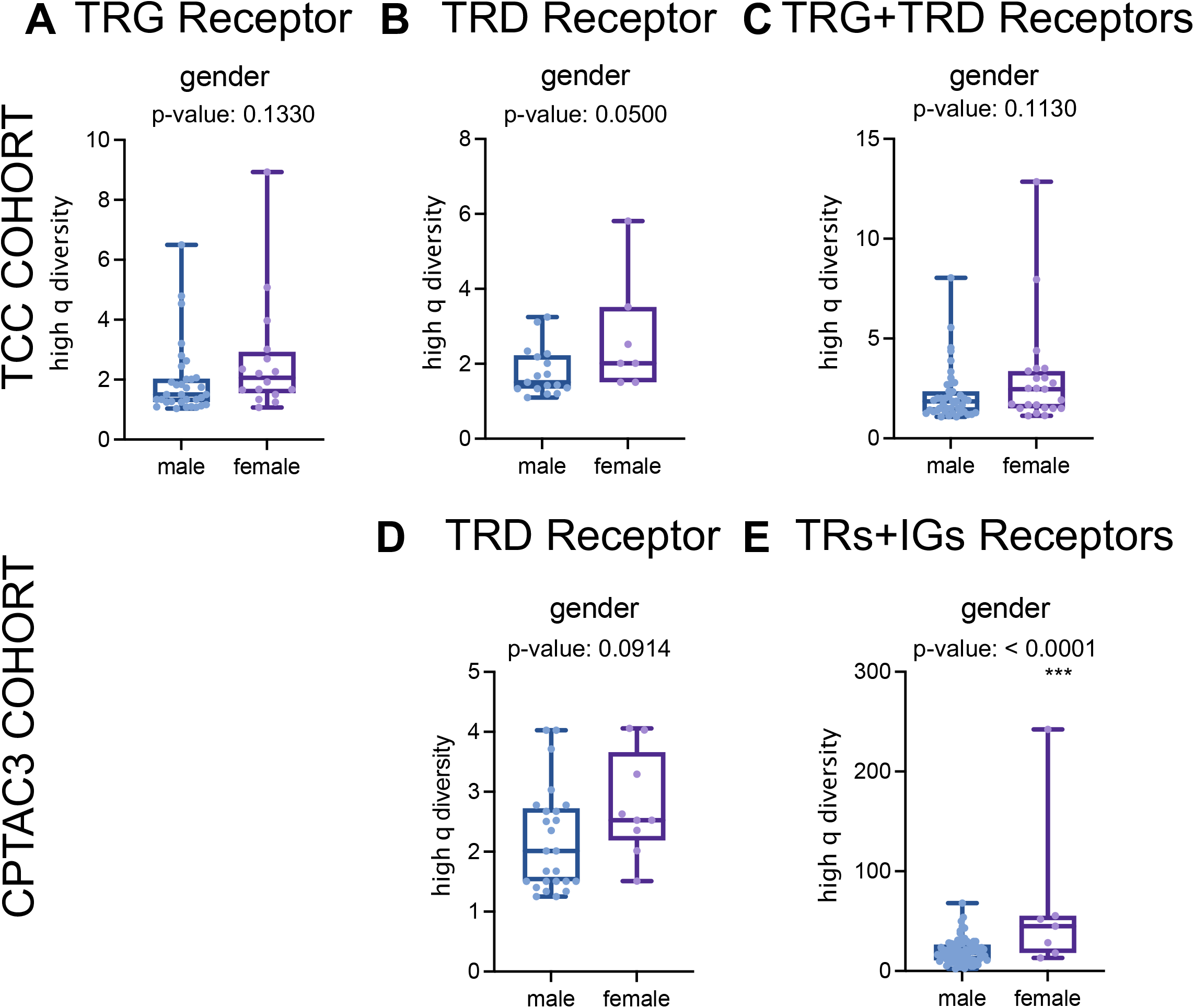
Gender-based diversity differences trends exist in T-cell receptor evenness groupings in both the Moffitt TCC Cohort and the CPTAC-3 Cohort. (A) T-cell receptor gamma (TRG) high q diversity was elevated in Moffitt TCC female patients compared to males (mean score in male patients was 1.963, mean score in female patients was 2.663; p-value: 0.1330). (B) T-cell receptor delta (TRD) high q diversity was elevated in Moffitt TCC female patients compared to males (mean score in male patients was 1.791, mean score in female patients was 2.699; p-value: 0.0500). (C) Combination TRG and TRD high q diversity was also elevated in Moffitt TCC female patients (mean score in male patients was 2.195, mean score in female patients was 2.977; p-value: 0.1130). These findings in the Moffitt TCC Cohort were supported in the CPTAC-3 Cohort. (D) TRD high q diversity was elevated in CPTAC-3 female patients compared to males (mean score in male patients was 2.183, mean score in female patients was 2.769; p-value: 0.0914). (E) Combination TRG and TRD high q diversity was also elevated in CPTAC-3 female patients (mean score in male patients was 19.61, mean score in female patients was 64.90; p-value: < 0.0001; excluding the outstanding female score of 242 reduces the mean female score to 35.35 with a p-value of 0.0042 significant difference compared to the male scores). Unpaired t-tests were used to compare categorical data. p-value significance represented by * < 0.05, ** < 0.01, *** < 0.001

**Supplemental Figure S20:**
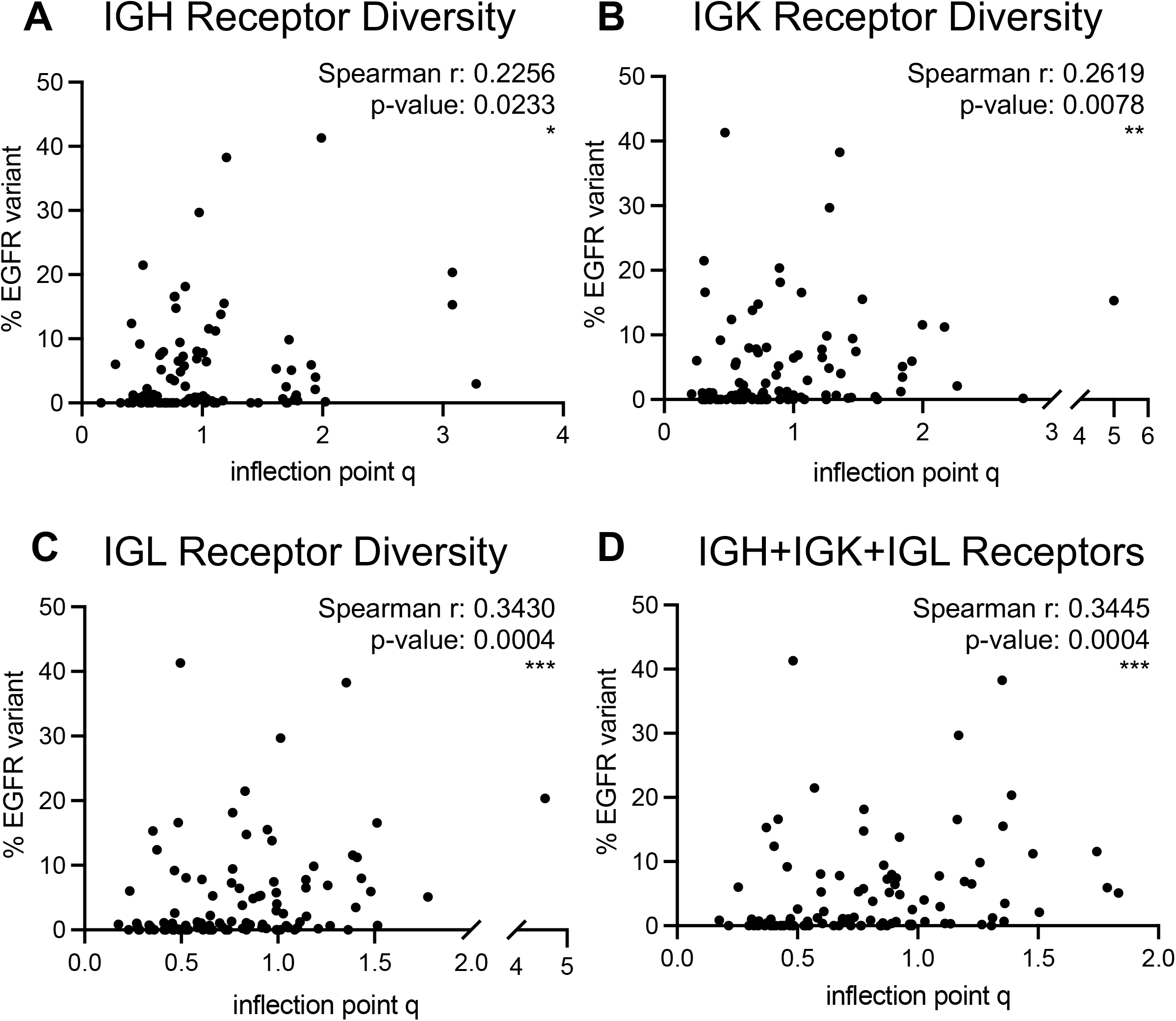
B-cell receptor inflection point q diversity may be a surrogate for the proportion of a renal cell carcinoma tumor with an EGFR variant. (A) IGH receptor inflection point q measures versus % EGFR variant reflected a significant weak correlation (Spearman r: 0.2256; p-value: 0.0233). (B) IGK receptor inflection point q measures versus % EGFR variant reflected a significant weak correlation (Spearman r: 0.2619; p-value: 0.0078). (C) IGL receptor inflection point q measures versus % EGFR variant reflected a significant weak correlation (Spearman r: 0.3430; p-value: 0.0004). (D) Full combination of B-cell receptor inflection point q measures versus % EGFR variant reflected a significant weak correlation (Spearman r: 0.3445; p-value: 0.0004). p-value significance represented by * < 0.05, ** < 0.01, *** < 0.001

**Supplemental Figure S21:**
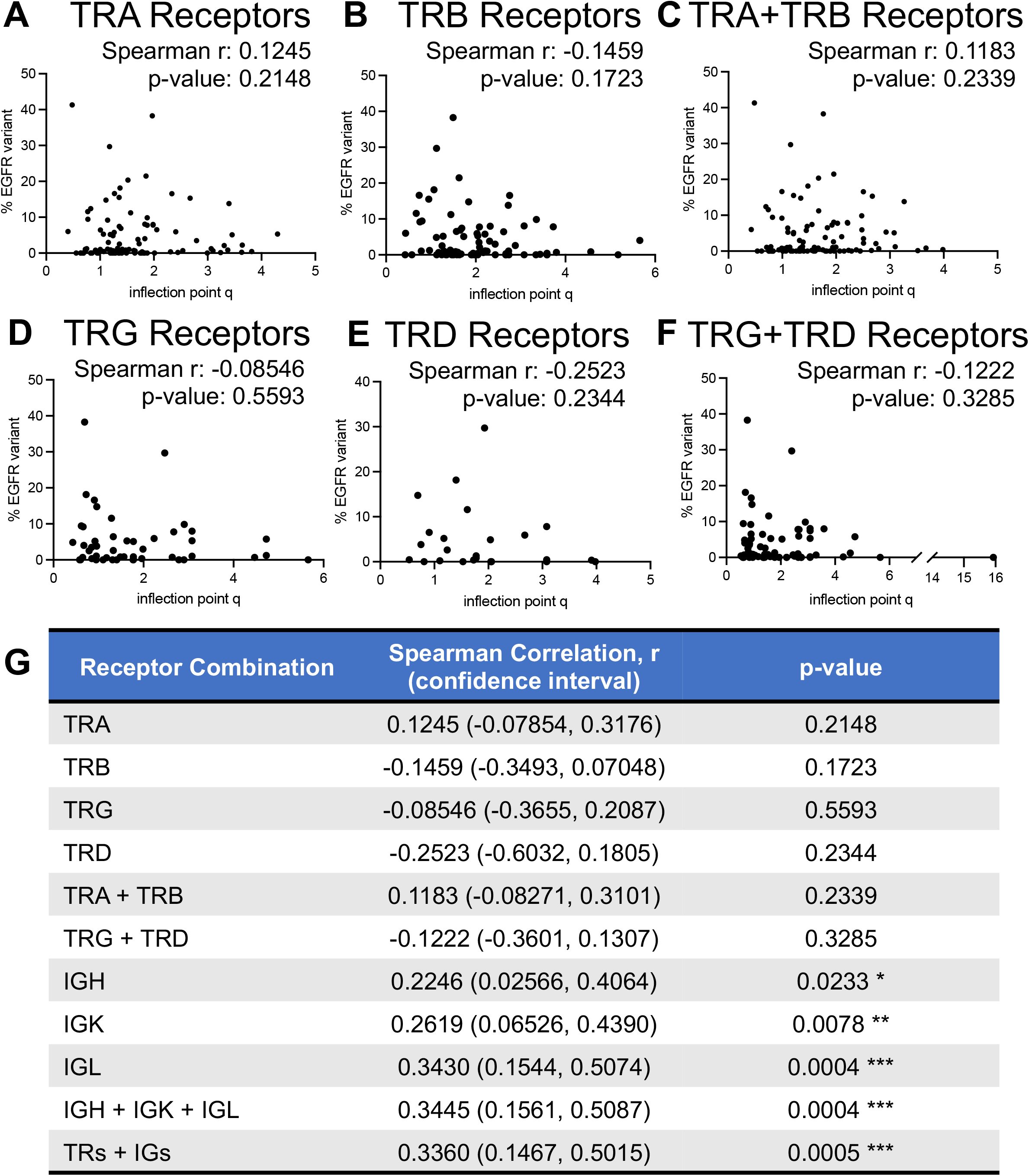
T-cell receptor inflection point q diversity does not significantly correlate with % EGFR variant in the Moffitt TCC Cohort. No significant correlations were evaluated for inflection point q measures versus % EGFR variant in recovered (A) TRA receptors, (B) TRB receptors, (C) combined TRA and TRB receptors, (D) TRG receptors, (E) TRD receptors, or (F) combined TRG and TRD receptors. (G) Summary table of correlation statistics for all receptor combinations. p-value significance represented by * < 0.05, ** < 0.01, *** < 0.001

**Supplemental Figure S22:**
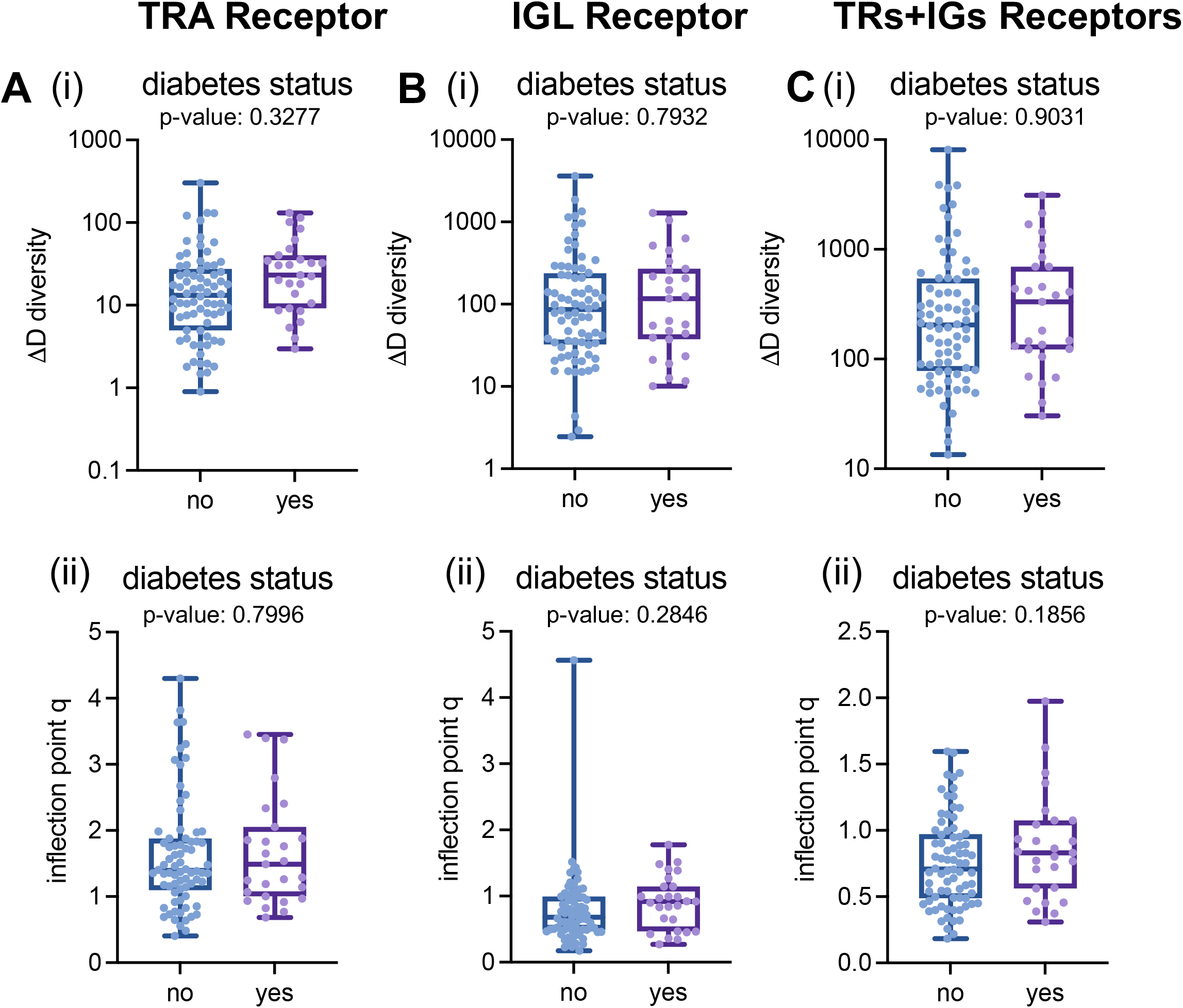
No associations with diabetes status and CDR3 diversity metrics in the Moffitt TCC Cohort. No significant associations were evaluated for ΔD diversity and inflection point q measures versus diabetes status in recovered (A) TRA recoveries, (B) IGL recoveries, (C) total (TRs+IGs) recoveries. (A) TRA receptors had (i) ΔD diversity had a mean score of 35.19 in individual with diabetes and mean score of 26.11 in those who did not have diabetes (p-value: 0.3277) and (ii) inflection point q had a mean score of 1.671 in individuals with diabetes and mean score of 1.623 in those who did not have diabetes (p-value: 0.7996). (B) IGL receptor had (i) ΔD diversity had a mean score of 231.2 in individual with diabetes and mean score of 259.2 in those who did not have diabetes (p-value: 0.7932) and (ii) inflection point q had a mean score of 0.9042 in individuals with diabetes and mean score of 0.7808 in those who did not have diabetes (p-value: 0.2846). (C) Total T-cell and B-cell receptors had (i) ΔD diversity had a mean score of 574.2 in individual with diabetes and mean score of 504.4 in those who did not have diabetes (p-value: 0.9031) and (ii) inflection point q had a mean score of 0.8662 in individuals with diabetes and mean score of 0.7601 in those who did not have diabetes (p-value: 0.1856). Unpaired t-tests were used for these analysis with p-value significance represented by * < 0.05, ** < 0.01, *** < 0.001

**Supplemental Figure S23:**
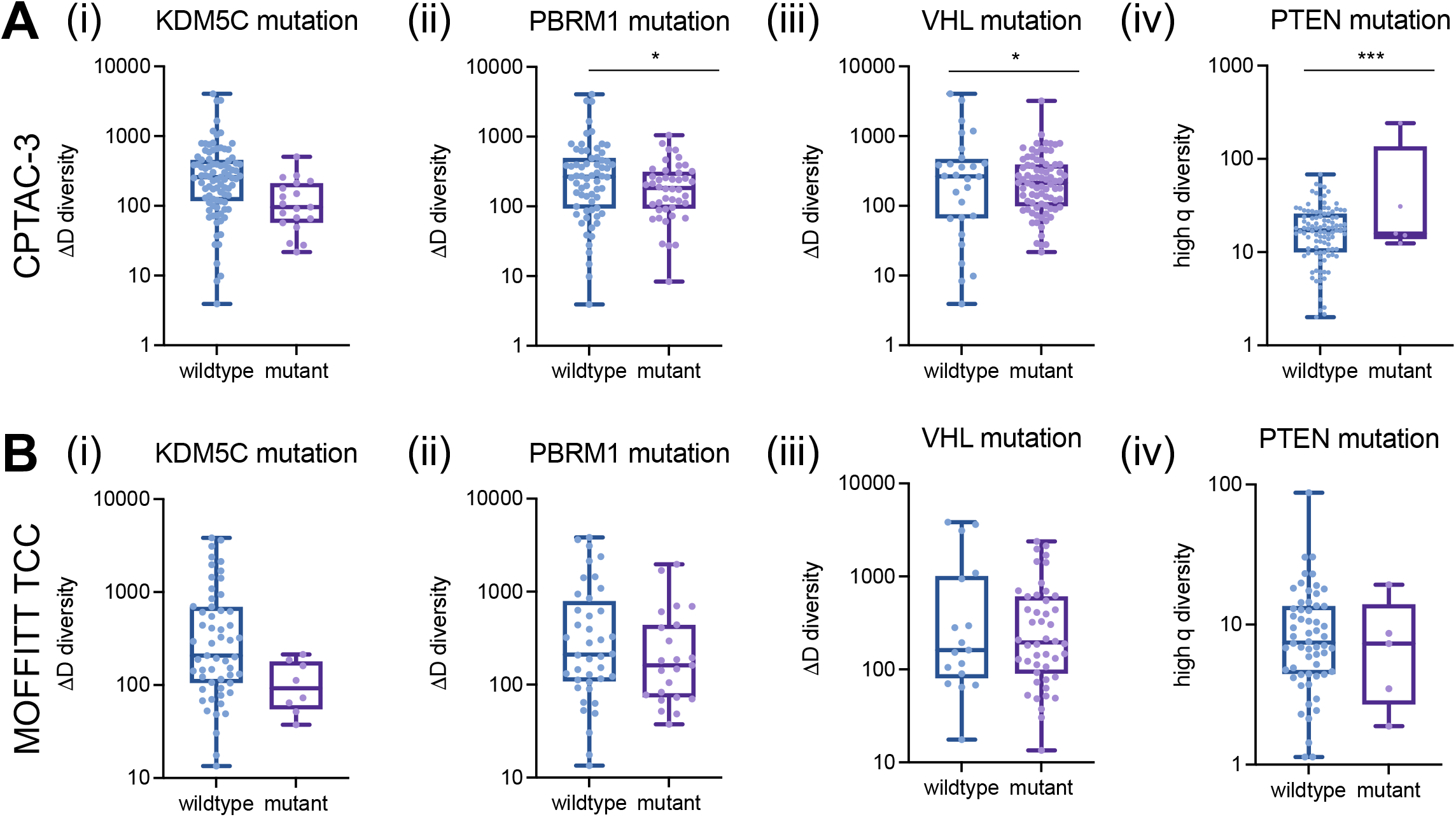
Total (TRs+IGs) recovery and diversity metric associations over the mutational landscape in clear cell renal cell carcinoma. (A) Total (TRs+IGs) recoveries had reduced richness in CPTAC-3 patients with (i) KDM5C mutations (mean score of wildtype was 431.1 and mutant was 138.0; p-value: 0.0515), (ii) PBRM1 mutations (mean score of wildtype was 473.7 and mutant was 240.7; p-value: 0.0448), and (iii) VHL mutations (mean score of wildtype was 575.9 and mutant was 313.8; p-value: 0.0446) and increased evenness in patients with (iv) PTEN mutations (mean score of wildtype was 19.14 and mutant was 63.30; p-value: <0.0001). (B) Total recoveries had reduced richness in Moffitt TCC patients with (i) KDM5C mutations (mean score of wildtype was 627.6 and mutant was 112.4; p-value: 0.1128), (ii) PBRM1 mutations (mean score of wildtype was 674.6 and mutant was 366.6; p-value: 0.1716), and (iii) VHL mutations (mean score of wildtype was 835.1 and mutant was 461.3; p-value: 0.1253) and increased evenness in patients with (iv) PTEN mutations (mean score of wildtype was 10.92 and mutant was 8.121; p-value: 0.6183). Unpaired t-tests were used to compare two group data and ANOVA was used to compare grade, three group data. p-value significance represented by * < 0.05, ** < 0.01, *** < 0.001

